# A wealth index based on two-component polychoric principal component analysis reduces urban bias and improves socioeconomic classification in low- and middle-income country surveys: a validation study using LSMS surveys

**DOI:** 10.64898/2026.06.01.26354245

**Authors:** Luis P. Vidaletti, Anderson Moreira Aristides dos Santos, Franciele Hellwig, Aluísio J. D. Barros

## Abstract

**Background:** The traditional PCA-based wealth index used in DHS and MICS surveys suffers from urban bias, distorting estimates of health inequality. We compared the traditional index (PEAR1) with an alternative two-component polychoric PCA index (POLY2) using annual expenditure from 12 LSMS surveys as the gold standard to determine which provides more accurate SEP measures for equitable policy targeting.

**Methods:** We compared the traditional wealth index (PEAR1) with a two-component polychoric PCA approach (POLY2) using 12 LSMS (Living Standards Measurement Study) surveys (2015-2022) from 12 African countries. Annual household consumption expenditure was the gold standard. We assessed agreement using weighted Cohen’s kappa and validated against education (proportion of households with secondary or higher education) using concentration (CIX) and slope (SII) indices of inequality.

**Results:** The POLY2 index showed higher agreement with expenditure quintiles (average national weighted kappa = 43.3%) than the PEAR1 index (35.1%), with notable improvements in urban (43.5% vs. 27.5%) and rural (35.3% vs. 22.4%) areas. POLY2 also attenuated extreme household distributions observed in PEAR1. Education validation showed that POLY2 produced intermediate inequality gradients between the flatter expenditure-based gradient and the steeper PEAR1-based gradient.

**Conclusion:** The POLY2 wealth index is superior to the traditional index, reducing urban-rural bias and providing more accurate socioeconomic classifications. Its adoption in large-scale surveys such as DHS and MICS is recommended to improve equitable monitoring of health inequalities in low- and middle-income countries.

## 1. Introduction

Socioeconomic position (SEP) is an important measure for classifying individuals or populations based on economic and social factors such as income, education, consumer goods, and occupation. It is widely recognized as one of the main social determinants of health, impacting mortality, disease outcomes, nutritional status, and access to health services.(Ramesh Masthi et al., 2013) There is a strong relationship between family wealth and maternal and child health outcomes, with marked pro-rich inequalities.(Baker et al., 2019; A. J. Barros & Victora, 2013; Cata-Preta et al., 2021; Cui & Chang, 2021; Filmer & Pritchett, 1998; Poirier et al., 2020; Wong et al., 2017) It is in low- and middle-income countries that we find the greatest reproductive, maternal, newborn, and child health (RMNCH) inequalities, due to a combination of structural, systemic, and individual factors.(A. J. D. Barros et al., 2020)

Large-scale household surveys such as the Demographic and Health Surveys (DHS) and Multiple Indicator Cluster Surveys (MICS) are fundamental for monitoring health inequalities in low- and middle-income countries (LMICs), as they collect comparable data on health indicators and key equity dimensions across most LMICs. However, these surveys do not collect expenditure or income data.

The proposed alternative for SEP classification was to create a wealth index using principal component analysis (PCA), based on data on goods and assets and some household characteristics (such as drinking water source, toilet type, lighting, among others). This approach, proposed by Filmer & Pritchett, was first published in 1998 as a World Bank technical report and in 2001 as a journal article.(Filmer & Pritchett, 2001)

Since then, the PCA-based wealth index has been widely used as a proxy for SEP in DHS and MICS surveys, as well as in other national health surveys. Although it solved an important problem, this approach has limitations. The main one is urban bias, as well-documented in the literature.(Dirksen et al., 2022; Martel et al., 2021; Poirier et al., 2020; Xie et al., 2023) The indicator overestimates the wealth of urban households relative to rural households, distorting inequality measurements.(Martel et al., 2021; Poirier et al., 2020) In some extreme cases, such as in Zimbabwe in 2014 (MICS) and 2015 (DHS), no urban household is classified in the first wealth quintile (Q1, the poorest 20%) or Q2. In Ukraine (MICS, 2005), Colombia (DHS, 2010), and Georgia (MICS, 2018), no rural household is classified in Q5 (the richest 20%) or Q4 (the second richest quintile). (https://dhsprogram.com/, https://mics.unicef.org/surveys)

In 2008, Shea O. Rutstein proposed a methodological update to the wealth index, including variables relevant to rural areas, such as the type and number of livestock and the size of agricultural land.(Shea O. Rutstein, 2008) The wealth index incorporated these variables through a more complex estimation process. This new approach improved the wealth estimation in rural areas. However, despite the more complex methodology that assigns separate scores to urban and rural areas and then combines them, it did not eliminate the urban bias. In contrast, Poirier et al.(Poirier et al., 2020) argue that the supposed urban bias is not a misclassification but rather an accurate reflection of the relative prosperity of urban families. While we acknowledge that urban areas are generally wealthier, the systematic overestimation of urban wealth — as shown by extreme cases in which no urban household falls into the poorest quintile — suggests that the index amplifies urban-rural differences beyond what is justified by actual prosperity. The persistence of this bias, even after methodological updates, indicates that the problem may be statistical and conceptual in nature. The purpose of a wealth index is to rank households for inequality analyses. An index that systematically overestimates urban wealth and underestimates rural wealth distorts estimates of health inequality, typically inflating the apparent gap in health coverage across wealth quintiles.

To attenuate this bias, the use of PCA based on a polychoric correlation matrix rather than Pearson correlation, and the consideration of the first two components rather than just the first, were proposed.(Kolenikov & Angeles, 2009; Martel et al., 2021) The advantage of the polychoric correlation matrix is that it treats multi-categorical variables, such as house construction materials, as ordinal, avoiding the dichotomization used in the traditional PCA (Pearson correlation matrix with one component) approach and preserving the qualitative order of the materials. The use of the second component enhances information on rural households. Although previous studies(Howe et al., 2008; Martel et al., 2021; Poirier et al., 2021; Ward, 2014) have used polychoric PCA, none have performed validation across a wide range of variables and in multiple countries, making it difficult to assess whether there is a systematic advantage of polychoric PCA over traditional PCA.

Given the limited efforts in the literature so far, our aim is to improve and expand the comparison and validation of the two wealth-based indices. Thus, we aim to compare the household wealth obtained through the current DHS approach (PEAR1) with that using the first two components of a PCA based on a polychoric correlation matrix (POLY2). This comparison is based on how each of these two classifications compares to the classification obtained from household consumption using LSMS surveys. To extend the validation to countries with different income levels and urbanization rates, we selected 12 LSMS surveys with suitable data collected since 2015.

## 2. Methods

### 2.1. Data source

To compare asset-based socioeconomic classifications, it is important to use a benchmark classification that serves as the gold standard. LSMS surveys are a natural choice, as they typically include annual expenditure data, household consumption estimates, data on household goods, and other socioeconomic indicators.

The World Bank’s LSMS surveys offer several advantages: access to annual expenditure data (the gold standard of economics),(Howe et al., 2008; Poirier et al., 2020) which allows for testing hypotheses across a wide range of countries from 2015 onwards and different contexts, thus increasing the external validity of the conclusions, and provides a sufficient sample size that meets all the assumptions of a rigorous study design for stratified analyses and more accurate concordance analysis estimates. Furthermore, it is a collection of research studies on well-being and socioeconomic determinants, which has been fully supported by the World Bank since 1985.

We applied the following inclusion criteria: national surveys conducted from 2015 onwards (to ensure contemporary relevance), with data on annual consumption expenditure and the variables necessary for constructing wealth indices. When countries conducted multiple surveys during this period, we selected the most recent one. From the LSMS repository (165 surveys in total), we excluded 22 non-national or thematic surveys (by telephone, on COVID-19, forests, land and soil, agriculture, among others). Of the remaining 143 surveys, after applying the time limit (from 2015 onwards), 37 remained, from 14 countries. We excluded Cambodia (without annual expenditure data) and Nigeria (without rural variables), resulting in 12 surveys in the final analysis.

### 2.2. Construction of measures of socioeconomic position

#### 2.2.1. Annual expenditure

Annualized household consumption expenditure is available in the LSMS databases across 13 groups: food and beverages; alcohol and tobacco; clothing and footwear; housing and utilities; furniture and equipment; health; transport; information and communication; recreation; education; restaurants; insurance; and personal care. To obtain the final variable, we summed the values for each household within each group, following the standard procedure for constructing total household expenditure from LSMS data. Total household expenditure quintiles were created by dividing households into five groups of equal population size, weighting by household size, and using sample weights.(Shea O. Rutstein, 2008)

#### 2.2.2. Traditional wealth index

The traditional wealth index was applied to the selected surveys, following the same steps described in Shea Rutstein’s article(Shea O. Rutstein, 2008) and detailed in the supplementary material. Basically, variables are created for all households and, except for the variables “number of people per sleeping room” and “agricultural land size”, which are entered as continuous, all other variables are entered into the analysis as binary, including each categorical variable.

First, a PCA based on Pearson’s correlation matrix is performed using all households, excluding livestock variables and agricultural land size, and the scores for the first principal component are extracted. Then, a PCA is performed for urban households only, and the scores for the first component are extracted. Finally, the scores for the first component are obtained from a PCA that includes only rural households and livestock variables, as well as agricultural land size.

The final adjusted scores are obtained by calculating predicted values from two linear regression models, one for each area, with PCA scores as the outcome and the area-specific PCA scores as predictors. The adjusted scores are combined into a single database, and wealth quintiles are created using household sample weights and the number of household members to form equal-sized groups.

#### 2.2.3. Polychoric wealth index with two components

According to Martel et al.,(Martel et al., 2021) variables that are originally yes/no remain as binary variables. Categorical household characteristic variables (e.g., construction materials, cooking fuel, lighting, garbage collection, etc.) are recategorized and treated as ordinal variables, with three or four categories (from worst to best), as shown in Table 1 and supplementary material (Tables SM7-SM21).

**Table 1.**
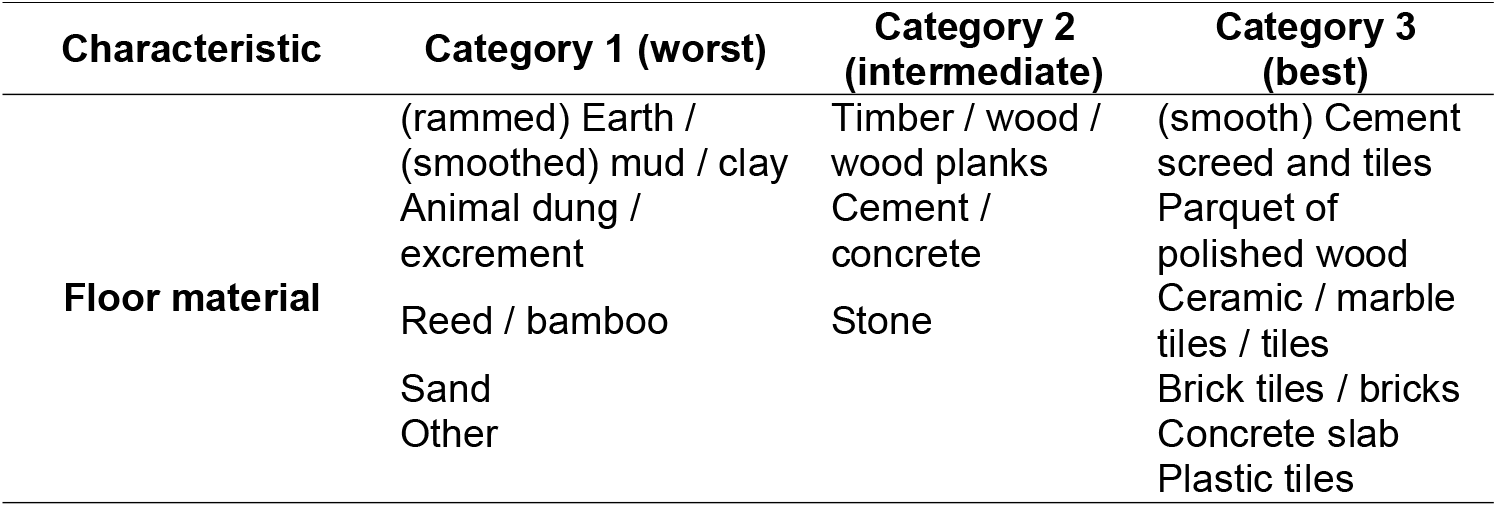
Classification of floor material categories into three ordinal levels (worst to best)

| Characteristic | Category 1 (worst) | Category 2 (intermediate) | Category 3 (best) |
| --- | --- | --- | --- |
| Floor material | (rammed) Earth / (smoothed) mud / clay | Timber / wood / wood planks | (smooth) Cement screed and tiles |
|  | Animal dung / excrement | Cement / concrete | Parquet of polished wood |
|  | Reed / bamboo | Stone | Ceramic / marble tiles / tiles |
|  | Sand |  | Brick tiles / bricks |
|  | Other |  | Concrete slab |
|  |  |  | Plastic tiles |

Instead of using PCA based on the Pearson (product-moment) correlation matrix, we used the polychoric correlation matrix, which is more appropriate for ordinal variables. The polychoric correlation matrix respects the ordering of categories(Kolenikov & Angeles, 2009; Olsson, 1979) (e.g., natural floor is inferior to rudimentary floor, which in turn is inferior to finished floor). In polychoric PCA, distances between categories are not assumed to be equal (e.g., between “earth floor”, “wood planks”, and “tiles”), but are estimated from the underlying normal distribution of responses, which better reflects relationships between ordinal variables.(Holgado-Tello et al., 2010) Furthermore, the PEAR1 approach almost always estimates negative loadings for rural variables.

In this approach, instead of using only the first component, we use the first two main components to compose the final wealth score. Typically, the first component reflects general aspects of wealth, with a greater focus on urban goods and assets.(Martel et al., 2021) The second component captures more aspects of rural wealth, assigning greater weight to area-specific variables.(Martel et al., 2021) The final score is simply the sum of the scores extracted from the first two components (a continuous latent variable), a strategy similar to summing the component vectors. With the final scores, households can be classified into quintiles (the most popular) or any other quantile group.

#### 2.2.4. Variables used in both PCA approaches

The survey questionnaires present similar lists of goods and assets variables, with some variations to reflect local characteristics and wealth levels. For example, in very poor areas, items such as beds and chairs are recorded, while in countries where these items are almost universal, they are not. Tables SM1 to SM6 present the lists of variables used in the analysis, in addition to dichotomous variables: domestic servant, agricultural land, property ownership, sharing a toilet with members of other households, local business, livestock ownership (yes/no), and services such as electricity, internet, cable TV, and piped water.

For household characteristics included in all analyses (e.g., type of dwelling, wastewater disposal, drinking water source, etc.), Tables SM7-SM21 present the variable categories and the proposed orderings (supplementary material).

Continuous variables include the number of household members per sleeping room, the total area of agricultural land used by all household members, and the number of each livestock. Livestock included in this analysis includes chickens, donkeys, goats, sheep, pigs, and rabbits, among others.

### 2.3. Comparison between the different approaches

The statistical analysis is based on three parts: first, an exploratory comparative analysis of the distributions by quintiles of annual expenditure and quintiles of wealth estimated by the PEAR1 and POLY2 for urban, rural, large urban center, and 100% rural districts; second, a comparison of the concordance analysis between the wealth quintiles estimated by the PEAR1 and POLY2 with the quintiles of annual expenditure; and finally, validation with an indicator of percentage of households with at least one resident with secondary or higher education, and indicator that is known to be strongly linked to socioeconomic level. While investigating the association between the SEP indicators and a health RMNCH outcome would also be interesting, LSMS surveys have data on health indicators for only a small portion of the total sample.

The initial descriptive analyses aim to characterize the research. Following the approach of Martel et al.(Martel et al., 2021), we initially compared the distributions of households by wealth quintiles, according to each approach (PEAR1 and POLY2), with the distributions of expenditure quintiles, stratified by urban and rural areas, in a large urban center in each country and in entirely rural districts.

The main analysis estimates the agreement between the PEAR1 and POLY2 quintiles and the annual expenditure quintiles. We measured observed agreement, which measures the degree of agreement between the rankings of two approaches assessing the same phenomenon, and the weighted Cohen’s kappa coefficient, which measures agreement beyond what is expected by chance. These agreements were tested on the total (national) sample and on the two residential areas (urban and rural), separately for each country, to assess the reduction of urban bias.

Finally, for each country and each approach (PEAR1, AEXP, POLY2), we stratified the proportion of households with at least one member with secondary or higher education by quintile, in the total sample (national level) and separately in urban and rural areas. Education is a key component of socioeconomic status and, in some contexts, is less susceptible to measurement errors than expenditure itself. We expect a monotonic positive gradient (higher education in the wealthiest quintiles). A flatter or non-monotonic gradient would indicate poor performance of the wealth index. All analyses accounted for the sample weights derived from each survey’s complex design and the number of household members.

To explore wealth-related inequalities in education, the following complex summary measures, which account for the entire socioeconomic distribution, are used: the concentration index (CIX) for relative measures and the slope index of inequality (SII) for absolute measures, with level of education (secondary or higher) as the dependent variable. Both measures range from −1 to 1 (in figures and tables, they are presented multiplied by 100), with positive values indicating that the dependent variable (education) is concentrated among the wealthy and negative values suggesting a pro-poor inequality.(A. J. Barros & Victora, 2013) The CIX is calculated similarly to the Gini index, with a graphical representation of the cumulative distribution of the indicator across wealth quantiles (quintiles in the analysis). The resulting curve is compared to the diagonal line indicating zero inequality. The SII is the absolute difference, in percentage points, between the poorest and richest extremes of the distribution, adjusted by logistic regression.(Wehrmeister et al., 2020)

## 3. Results

### 3.1. Description of the surveys

Table 2 presents the 12 surveys used in the analysis. Except for Malawi and Uganda (2019-2020), all other surveys collected data during the same period, 2021-2022. The average sample size observed was approximately 6,800 households. Of these, about 87% had at least one household member who answered the question about the highest level of education achieved. The average rural population in these countries was approximately 64%, ranging from 45% in the Côte d’Ivoire to 84% in Niger. The number of variables used in each analysis averaged 38 dummy variables, 10 categorical variables, and 9 continuous variables, resulting in 57 variables per country (on average).

**Table 2.**
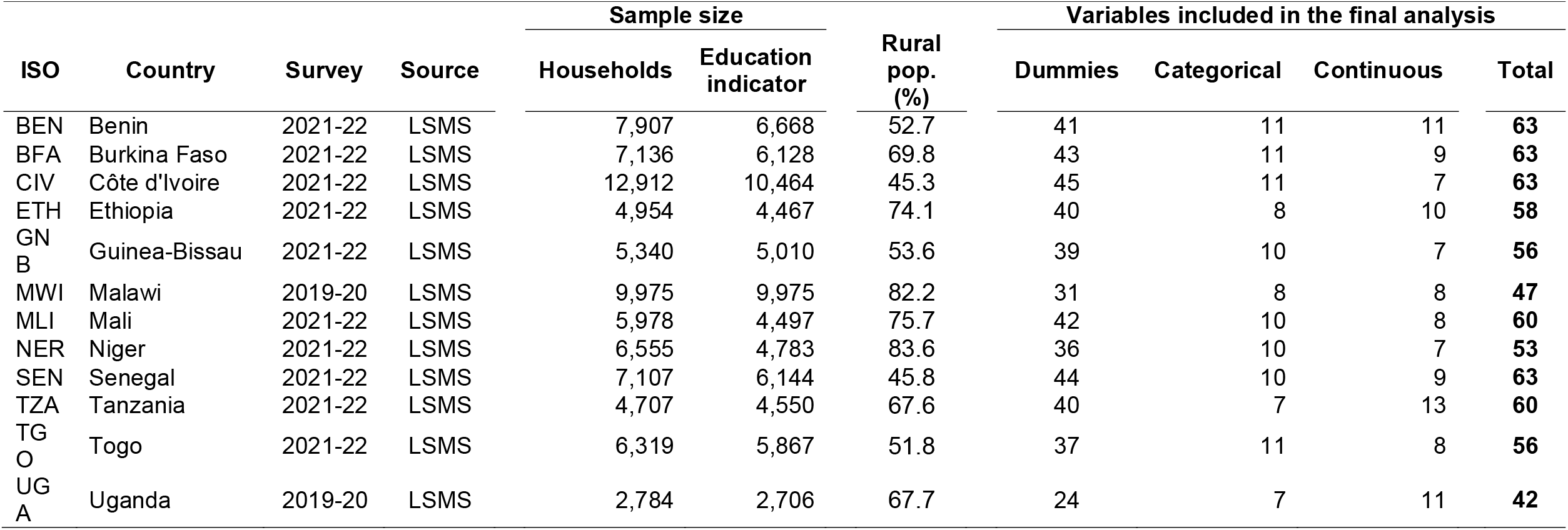
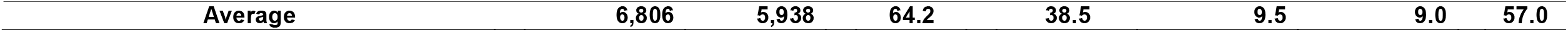
List of countries included, year of the survey, sample size, percentage of the rural population, and number of variables used.

### 3.2. Exploratory analysis of households’ distributions

In Figure 1, we observe that the percentage distributions of households estimated by POLY2 closely approximate annual expenditures, whereas those estimated by PEAR1 in Tanzania (for instance) do not. Analyzing the distribution of quintiles generated by PEAR1 in the urban area, 60% of households were classified as the richest. Estimating by annual expenditure, this percentage is only 37%, and by POLY2, it is 44%. In Dar es Salaam region, PEAR1 classified 84% of households in the richest quintile and 0% in the three poorest quintiles. For expenditure and POLY2, we observed 46% and 54% in the richest quintile, respectively. And for the three poorest quintiles, we found 26% (AEXP) and 17% (POLY2). In the rural area, the percentages in the richest quintile are 5% for PEAR1, 14% for annual expenditure, and 11% for POLY2. The same approximation of POLY2 to AEXP is also observed in the distributions of 100% rural districts.

**Figure 1.**
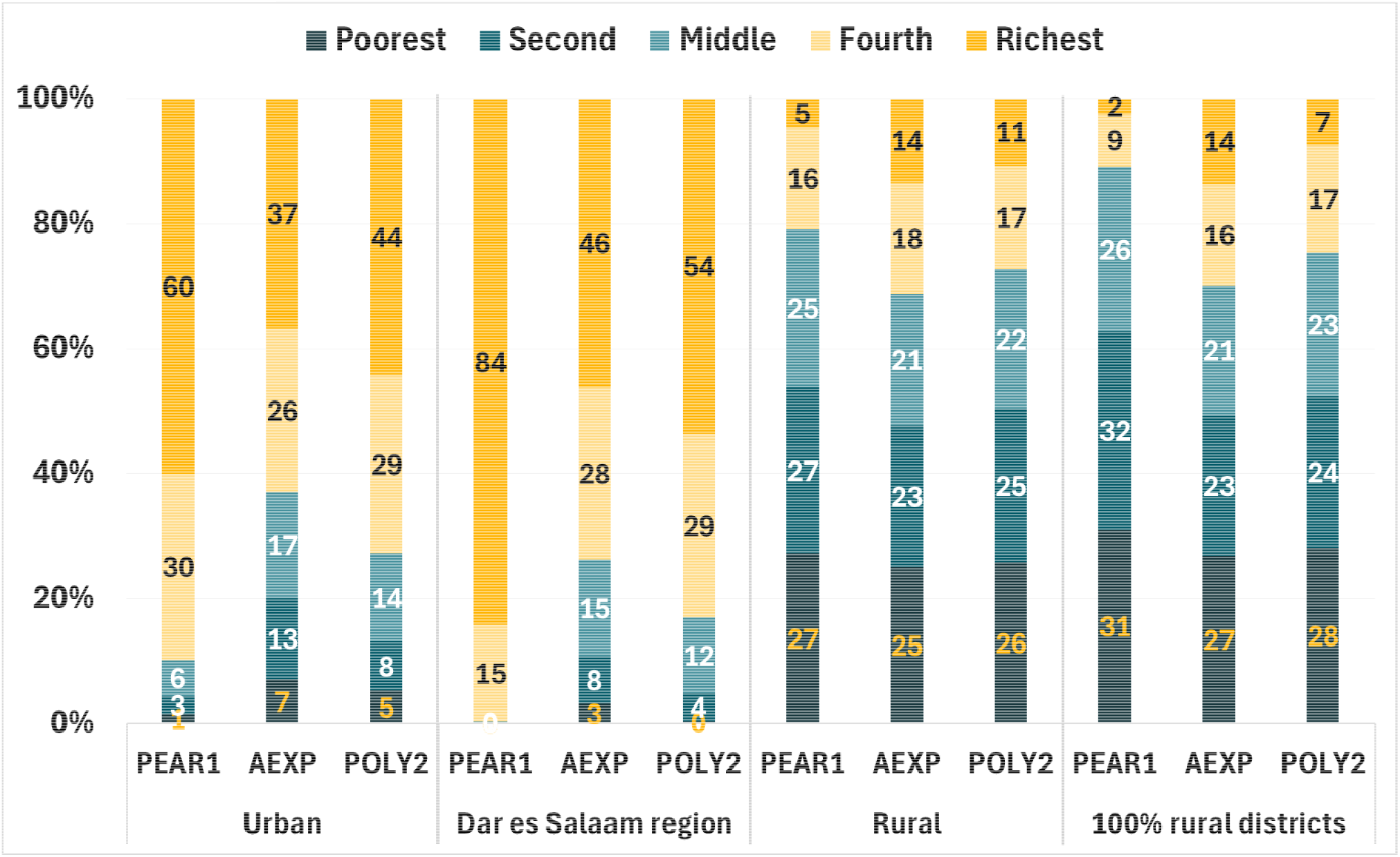
Percentage distributions of urban, rural, main urban center, and 100% rural district households by quintile for each approach – Tanzania (2021 LSMS)

### 3.3. Agreement between wealth indices and expenditures

We can observe in Table 3 that, for more than half (7 out of 12) of the countries, both for observed agreement and for Cohen’s kappa coefficients (Kappa values are also presented as percentages), at the national level and for urban and rural areas, the agreement between household quintiles classified by POLY2 shows greater agreement with household quintiles classified by AEXP than between household quintiles classified by PEAR1 and annual expenditure quintiles. In Ethiopia, Guinea-Bissau, Mali, and Niger, the differences in agreement are not significant in some cases. Only Malawi showed greater agreement between PEAR1 and AEXP than between POLY2 and annual expenditure. The weighted-average kappa value for agreement beyond expectations, estimated between the POLY2 approach and annual expenditure across all countries, was 43.3% (national). This is substantially higher than the 35.1% observed between the PEAR1 and AEXP approaches. This improvement was even more pronounced in urban (43.5% vs. 27.5%) and rural areas (35.3% vs. 22.4%). Confidence intervals are available in the supplementary material—in Tables SM22-SM24.

**Table 3.**
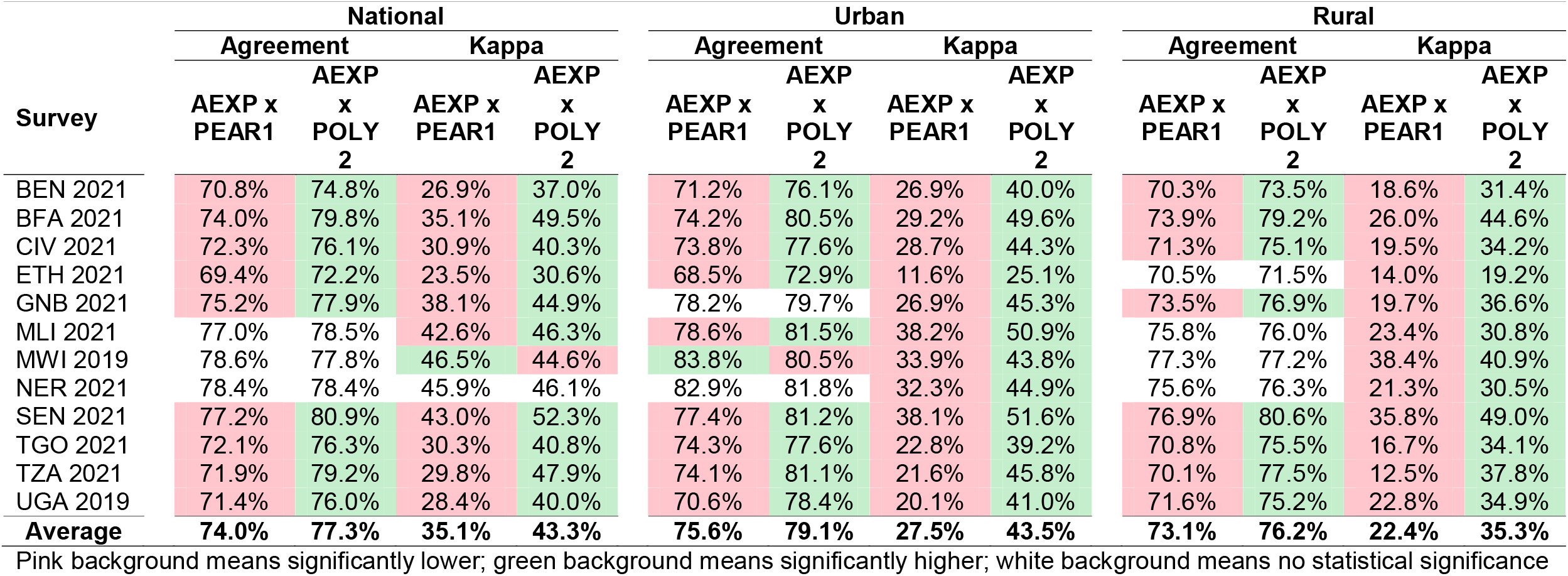
Comparison of approaches regarding observed agreement and the Kappa coefficient at the national, urban, and rural levels.

### 3.4. Validation with schooling

In the analysis of the equiplots, in Figure 2 and in the supplementary material (Figures SM12-SM22), we estimated graphs showing estimates of the proportion of households with at least one member with secondary or higher education, by quintile, based on the annual expenditure and wealth-estimated approaches of the PEAR1 and POLY2 at the national, urban, and rural levels. We also estimated summary measures of relative inequality (CIX — Concentration Index) and absolute inequality (SII — Inequality Slope Index) in Tables SM25-SM27. We observed that, in more than half of the cases, the inequality estimated by the PEAR1 is the highest, while the inequality by quintile is lowest for annual expenditure, leaving POLY2 as an intermediate level of inequality. Similarly, when observing the CIX and SII values, in more than half of the cases, the order of inequalities is the same – higher inequality in the PEAR1 and lower in annual expenditure.

**Figure 2.**
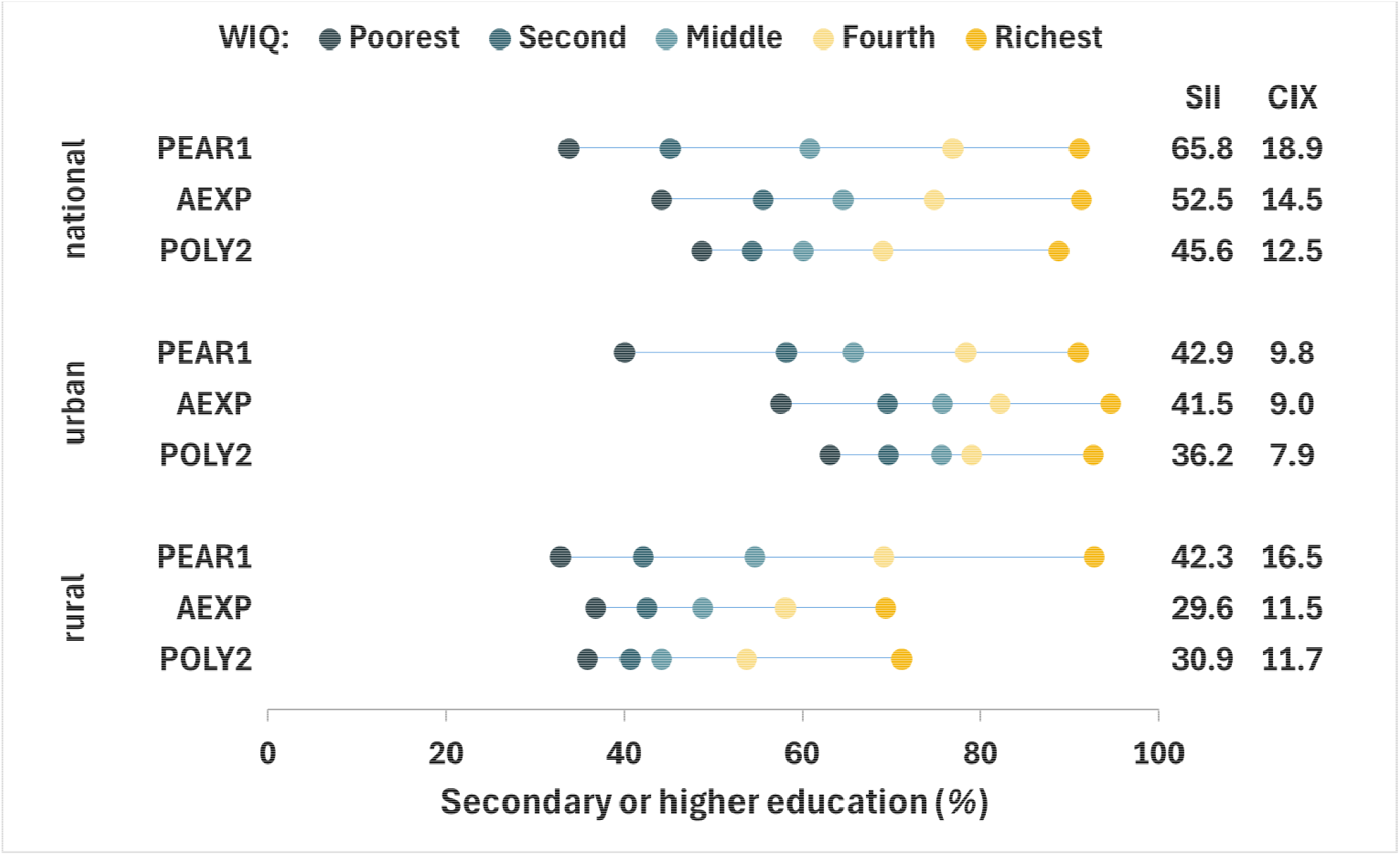
Proportion of households with at least one member with secondary or higher education by wealth quintile, approach, and geographic area – Côte d’Ivoire (2021 LSMS)

On average, the SII of the TRPCA approach was 6% higher than that of the POLY2 approach and 8% higher than that of annual expenses. Similar patterns were observed in the concentration index, with even greater differences – the CIXs of the PEAR1 quintiles are, on average, 14% higher than those obtained by POLY2 and 16% higher than the CIXs of annual expenses.

Considering only urban and rural strata, there is no significant difference between the SIIs and the CIXs obtained by PEAR1 and POLY2 in urban areas (−5% and 0.3%, respectively). However, at the national level and in rural areas, the differences between the SII of the quintiles obtained by PEAR1 and those obtained by POLY2 are, on average, about 11% higher, and those between the SII of the quintiles obtained by PEAR1 and the SII of annual expenditures are about 15% higher. Meanwhile, the CII of inequalities between quintiles estimated by PEAR1 are, on average, about 16% higher than those obtained by POLY2 and 19% higher than those of annual expenditures.

We observed that the poorest quintile generated by the PEAR1 in urban areas has a percentage of people with secondary or higher education much lower than those of the poorest quintiles estimated by POLY2 or by annual expenditures. Conversely, the richest quintile in rural areas, estimated by the PEAR1, has a percentage of people with secondary or higher education much higher those of the richest quintiles estimated by POLY2 and by annual expenditures.

## 4. Discussion

In summary, the main conclusion is that the wealth index estimated by POLY2 shows a household distribution more similar to that of annual expenditures in urban areas, large urban centers, and 100% rural districts. In almost all countries (11 out of 12), the distribution of households by quintile is more similar between the POLY2 approach and annual expenditures than between the PEAR1 and expenditures. This improvement was more pronounced in large urban centers and in the country’s urban areas, largely because richer households, as estimated by the PEAR1, were over-represented, leading poorer households to be classified as richer. The distributions estimated by the POLY2 approach approximate the distributions of annual expenditure, suggesting an effective reduction in urban bias. Only Malawi presented a different interpretation, perhaps because it is a country with more than 80% of its population living in rural areas/or because the research was conducted in 2019 (unlike most surveys, which are from 2021), or because fewer asset variables were included (31 vs. 38 average).

Our results confirm the findings of Martel et al.(Martel et al., 2021) and expand the dataset from a single country (Mozambique) to a diverse set of 12 nations in the POLY2 approach. Furthermore, by validating the POLY2 approach in the education context, we show that the estimated wealth index not only better fits expenditures but also provides a more plausible gradient for another key social determinant. Our study complements Mohanty’s study, which found evidence of overestimation among the urban poor in healthcare utilization, by providing broader validation of the POLY2 approach across multiple countries and by using educational outcomes as an additional benchmark.(Mohanty, 2009)

Validation with educational attainment confirmed that POLY2 more closely approximates the social gradient observed in the expenditure measure than PEAR1, especially in urban and rural areas, with emphasis on the extreme quintiles. A striking pattern emerged from our validation with education – according to PEAR1, the poorest urban households appeared substantially more disadvantaged relative to other urban quintiles (bottom inequality), while the richest rural households appeared substantially more advantaged relative to other rural quintiles (top inequality).(A. J. Barros & Victora, 2013) These patterns suggest that PEAR1 systematically exaggerates disadvantage among the urban poor and advantage among the rural rich — a “compression and expansion” bias consistent with the index’s tendency to overrepresent urban households in the richest quintiles and rural households in the poorest quintiles. POLY2’s intermediate position between expenditure (less unequal) and PEAR1 (more unequal) is theoretically desirable: a wealth index should detect meaningful inequalities without artificially amplifying them. This finding aligns with Martel et al., who observed similar attenuation of extreme distributions using the polychoric two-component index in Mozambique. In Niger, sample sizes for the poorest urban quintiles (N < 25) were too small to estimate inequality reliably in those strata.

Estimates of wealth indices obtained from a polychoric matrix with a second principal component make them more plausible. In the case of the polychoric matrix, respecting the ordinal nature of the family characteristic categories and using the second principal component, it better captures “rural wealth” (e.g., livestock and size of agricultural land), which is obscured by the urban assets of the first component—corroborating the articles by Ward(2014) and Martel et al.(2021) In rural areas, however, we found results distinct from those of Mohanty – our estimates for secondary or higher education showed a markedly gradient—the richer the family, the higher the estimates for secondary or higher education. For Mohanty, in rural areas, there is no observed gradient in the percentages of families using health services regarding their wealth.

The adoption of approaches such as POLY2 can lead to more accurate measurement of health inequalities, preventing rural populations from being systematically classified as poorer than they are and urban populations from being systematically classified as richer than they are.

Adopting this approach in public health studies can have direct implications for monitoring the SDGs (such as SDG 10 — reducing inequalities) and for allocating health resources, enabling more targeted and equitable interventions in low- and middle-income countries, especially in rural areas.

### 4.1. Limitations of the study

One of the main limitations of the study is its geographic scope: the sample of LSMS countries available for the selected period is entirely African, which may limit generalization to other regions, such as South Asia or Latin America. However, while we cannot claim that our findings generalize to other regions, we hypothesize that the POLY2 approach would also reduce urban bias in settings such as South Asia or Latin America, given its theoretical foundation. This remains an empirical question for future research. Our study is limited to African countries. Although the 12 countries vary in their UNICEF region (Ethiopia, Malawi, Tanzania, and Uganda in East and Southern Africa, and the others in West and Central Africa), World Bank income classification (Benin, Côte d’Ivoire, Senegal, and Tanzania as lower-middle-income countries, and the others as low-income countries), and rural population share (ranging from 45% in Côte d’Ivoire to 84% in Niger), this geographic restriction nevertheless limits generalization to other world regions such as South Asia or Latin America.

Using annual expenditures as a benchmark, while good practice, is also subject to reporting and, consequently, measurement errors. This is because it requires extensive questionnaires with detailed information over a considerable period, demanding memory and data recording.(Davila et al., 2022) This is especially true in populations with lower levels of education and a higher percentage of people living in rural areas, where families need less cash for many transactions. This can attentively at agreement estimates – that is, the actual agreement may be even greater than the observed agreement.

The availability and definitions of goods variables vary between surveys, slightly affecting comparability. However, this variability occurs only in some surveys. For example, seven of the 12 surveys (Benin, Burkina Faso, Côte d’Ivoire, Guinea-Bissau, Mali, Niger, and Senegal) investigated the same assets (45 in total). Togo surveyed 43 assets identical to those on the list in these seven surveys—except for guitars and pianos. Analyzing the choice of assets surveyed in other countries and using the selected range of these 45 assets as a basis, Tanzania stands out—it surveyed 37 of these assets, plus another 29. The same occurred in Ethiopia—it surveyed 18 of the 45 and ended up including another 20 assets in the questionnaire. Despite these differences, the POLY2 (polychoric with two components) principle remains consistent and can be adapted locally.

In estimating the wealth index using Pearson’s correlation matrix, some indicator variables are excluded from the analysis because they show no variation (zero standard deviation), typically due to a lack of cases(Shea Oscar Rutstein & Johnson, 2004) – or when only a single household has them – or even when only a single household in a residential area (urban/rural) has them. However, in the POLY2 process, a slightly larger number of variables are eliminated using multiple squared correlations (MSCs),(Martel et al., 2021) which corresponds to approximately 5.2% more. Nevertheless, these removed variables show a weak correlation with asset-based wealth.(Martel et al., 2021)

## 5. Conclusion

The wealth index estimated from the two-component polychoric correlation matrix is superior to the traditional wealth index—estimated in DHS and MICS surveys—by reducing urban bias and producing rankings that are more consistent with annual expenditures and educational gradients across 12 African countries.

This approach is a robust, feasible, and easily implemented alternative for national surveys such as the DHS and MICS. We recommend that research programs and researchers adopt the POLY2 approach for socioeconomic stratification.

Adopting the POLY2 approach would lead to more accurate measurements of health inequalities in low- and middle-income countries, preventing classifications that are systematically biased against rural areas (and towards urban areas), and enabling more targeted and equitable resource allocation.

## Data Availability

This study used de-identified, publicly available secondary data from LSMS surveys.

https://microdata.worldbank.org/index.php/catalog/lsms/?page=1&ps=15&repo=lsms

## Acknowledgements

The authors would like to thank Dr. Adriana Vieira Camerini for her constructive comments on an earlier draft of this manuscript.

