## Supplementary files for "A wealth index based on two-component polychoric principal component analysis reduces urban bias and improves socioeconomic classification in low- and middle-income country surveys: a validation study using LSMS surveys"

### Supplementary Material

#### Steps for constructing the standard Wealth Index

The methodology for replicating the DHS approach^1^ in constructing the wealth index, there are eight steps. Although the wealth index is based on household data, the first variables are created from the individual-level database. Step 1 consists of creating a variable indicating whether the household has a domestic servant. In Step 2, we create a variable indicating whether any household member owns land used for agriculture or whether the household's land is used for that purpose. Land size is also considered, with measurements standardized (e.g., square meters, hectares, acres).

Step 3 is generally the most labor-intensive, as it involves creating the most variables for the analysis. We begin by checking the number of rooms in the household used as bedrooms, then move to the variable that indicates whether the household is owned. After that, we move to categorical variables, transforming each variable's category into a dichotomous indicator variable. The categorical variables used are: drinking water source (surface water, spring, well, borehole, tap, piped water, etc.), toilet type (none, pit, latrine, flush, etc.), flooring, wall, and roofing materials (natural, rudimentary, finished, etc.), cooking fuel (firewood, charcoal, electricity, gas, etc.), lighting (paraffin, kerosene, generator, electricity), garbage collection (illegal dump, buried, burned, public collection), housing type (hut, room, shack, tent, traditional house, brick house, apartment, etc.), sewage (street or nature, hole in the ground, sewer, septic tank), as well as handwashing place (none, mobile place, fixed place) and kitchen type (none, inside the house, outside the house, etc.).

In this Step 3, we also check whether the toilet type is shared by members of other households, as well as electricity, air conditioning, water heater, ceiling fan, business on premises, piped water, internet, and cable TV. Step 4 is where we generate variables for goods and assets, such as: refrigerator, television, mobile phone, car/truck, computer, wardrobe, table, sofa, stove, chair, washing machine, iron, etc.

Step 5 consists of generating variables related to rural areas, such as livestock ownership. Both ownership (yes/no) and the total number of each type of livestock. This varies from country to country, but it generally includes rabbits, pigs, goats, chickens, sheep, and other poultry. Step 6 consists only of generating a variable indicating whether any household member has a bank account.

Steps 7 and 8 are very important and require attention. First, a variable is created for the number of household members per sleeping room, and subsequently, we check whether any indicator variables lack variation due to the absence of cases or because only a single household in the total sample possesses them. That variable is then excluded and does not enter the analysis, as is the verification of the frequency distribution of variables eligible for analysis by area of residence.

In the eighth and final step, principal component analyses are performed for all households, excluding the livestock variables and the agricultural land size variable, and the scores for the first principal component (PC1) are saved based on the Pearson correlation matrix. Then, two PCAs are run – one only for urban households and another for rural households, and the PC1 scores from each are saved.

Subsequently, two regression analyses are performed, with the outcome being the PCA scores for all households. For each area (urban/rural), a regression analysis is performed with the PCA scores (urban/rural) as predictors, using households from each area. Then, a regression analysis is performed with the PCA scores for all households as the outcome and, as predictors, the urban PCA scores (only for urban households), with alpha and beta saved. With this, a combined score variable is generated using the formula $gen combined=urban_{alfa}+urban_{beta}*urban_{score} if area=="urban"$ and subsequently replaced by $replace combined=rural\_alfa+rural\_beta*rura\_score if area=="rural"$. Finally, a quintile variable is created, weighted by multiplying the number of de jure household members by the household sampling weight.

Table SM1. Table of goods and assets used in the Malawi (2019 LSMS) survey

| **Asset variables** | |
| --- | --- |
| Mortar/pestle (mtondo) | Car |
| Bed | Minibus |
| Table | Lorry |
| Chair | Beer-brewing drum |
| Fan | Upholstered chair, sofa set |
| Air conditioner | Coffee table (for sitting room) |
| Radio ('wireless') | Cupboard, drawers, bureau |
| Tape or CD/DVD player; HiFi | Lantern (paraffin) |
| Television | Desk |
| VCR | Clock |
| Sewing machine | Iron (for pressing clothes) |
| Kerosene/paraffin stove | Computer equipment & accessories |
| Electric or gas stove; hot plate | Satellite dish |
| Refrigerator | Solar panel |
| Washing machine | Generator |
| Bicycle | Electric kettle |
| Motorcycle / Scooter | Radio with flash drive/micro-CD |

Table SM2. Table of goods and assets used in the Uganda (2019 LSMS) survey

| **Asset variables** | |
| --- | --- |
| Owner occupied House | Jewelry and Watches |
| Other Buildings | Mobile phone |
| Non-agricultural land | Computer - Desktop/Laptop/Tablet/iPad |
| Furniture/Furnishings | Internet access |
| Household Appliances e.g. Kettle, Flat iron, etc. | Other electronic equipment |
| Television | Other household assets e.g. lawn mowers, etc. |
| Cassette/DVD/CD | Other 1 (specify) |
| Generators | Other 2 (specify) |
| Solar panel/electric inverters | Cooker |
| Bicycle | Refrigerator |
| Motorcycle | Washing machine/Driers |
| Motor vehicle | Radio |
| Boat /Canoe | Home theatre / music system |
| Other Transport equipment | Fixed phone |

Table SM3. Table of goods and assets used in surveys in Benin, Burkina Faso, Côte d’Ivoire, Guinea-Bissau, Mali, Niger and Senegal (2021 LSMS)

| **Asset variables** | |
| --- | --- |
| Living room (Armchairs and coffee table) | Dining table (table + chairs) |
| Bed | Single mattress |
| Cabinets and other furniture | Carpet |
| Electric iron | Charcoal iron |
| Gas or electric cooker | Gas cylinder |
| Gas or electric stove (hob) | Microwave or electric oven |
| Improved fireplaces | Electric food processor (Moulinex) |
| Non-electric blender/Fruit press | Fridge |
| Freezer | Pedestal fan |
| Simple radio/Radio cassette | TV box |
| VCR/CD/DVD | Satellite dish / decoder |
| Washing machine, tumble dryer | Vacuum cleaner |
| Portable air conditioners/Split units | Lawnmower and other gardening equipment |
| Generator set | Personal car |
| Moped/Motorized bicycle, motorcycle | Bicycle/Racing bike |
| Camera | Camcorder |
| Hi-Fi system | Landline telephone |
| Mobile phone | Tablet |
| Computer | Printer/Fax |
| Video Camera | Dugout canoe and outboard motor (pleasure boats) |
| Hunting rifles | Guitar |
| Piano and other musical instruments | Building/House |
| Undeveloped land |  |

Table SM4. Table of goods and assets used in the Ethiopia (2021 LSMS) survey

| **Asset variables** | |
| --- | --- |
| Kerosene stove | Cylinder gas stove |
| Electric stove | Blanket/Gabi |
| Mattress and/or bed | Wristwatch/clock |
| Fixed line telephone | Radio/ tape recorder |
| Television | CD/VCD/DVD/Video Deck |
| Satellite dish | Sofa set |
| Bicycle | Motorcycle |
| Cart (hand pushed) | Cart (animal drawn)- for transporting people and goods |
| Sewing machine | Weaving equipment |
| Mitad-electric | Energy saving stove (lakech, mirt etc.) |
| Refrigerator | Private car |
| Jewels - Gold (in grams) | Jewels - Silver (in grams) |
| Wardrobe | Shelf for storing goods |
| Biogas stove (pit) | Water storage pit |
| Sickle (Machid) | Axe (Gejera) |
| Pick Axe (Geso) | Plough (traditional) |
| Plough (Modern) | Water pump |
| Solar device | Bajaj |
| Personal computer/Laptop |  |

Table SM5. Table of goods and assets used in the Tanzania (2021 LSMS) survey

| **Asset variables** | |
| --- | --- |
| Radio and radio cassette | Telephone (landline) |
| Telephone (mobile) | Refrigerator or freezer |
| Sewing machine | Television |
| Video / DVD | Chairs |
| Sofas | Tables |
| Watches | Beds |
| Cupboards, chest-of-drawers, boxes, wardrobes, bookcases | Lanterns |
| Computer | Cooking pots, cups, other kitchen utensils |
| Mosquito net | Iron (charcoal or electric) |
| Electric/Gas stove | Another stove |
| Water-heater | Record/Cassette player, tape recorder |
| Complete music system | Books (not schoolbooks) |
| Motor vehicles | Motorcycle |
| Bicycle | Carts |
| Animal-drawn cart | Boat/Canoe |
| Wheelbarrow | Livestock |
| Poultry | Outboard engine |
| Donkeys | Fields/Land |
| House(s) | Fan/Air conditioner |
| Dish antenna/Decoder | Hoes |
| Spraying machine | Water pumping set |
| Reapers | Tractor |
| Trailer for tractors etc. | Plough etc. |
| Harrow | Milking machine |
| Harvesting and threshing machine | Hand milling machine |
| Coffee pulping machine | Fertilizer distributor |
| Power tiller | Bajaj/Toyo |
| Guta | Incubator |

Table SM6. Table of goods and assets used in the Togo (2021 LSMS) survey

| **Asset variables** | |
| --- | --- |
| Living room (Armchairs and coffee table) | Dining table (table + chairs) |
| Bed | Single mattress |
| Cabinets and other furniture | Carpet |
| Electric iron | Charcoal iron |
| Gas or electric cooker | Gas cylinder |
| Gas or electric stove (hob) | Microwave or electric oven |
| Improved fireplaces | Electric food processor (Moulinex) |
| Non-electric blender/Fruit press | Fridge |
| Freezer | Pedestal fan |
| Simple radio/Radiocassette | TV box |
| VCR/CD/DVD | Satellite dish / decoder |
| Washing machine, tumble dryer | Vacuum cleaner |
| Portable air conditioners/Split units | Lawnmower and other gardening equipment |
| Generator set | Personal car |
| Moped/Motorized bicycle, motorcycle | Bicycle/Racing bike |
| Camera | Camcorder |
| Hi-Fi system | Landline telephone |
| Mobile phone | Tablet |
| Computer | Printer/Fax |
| Video Camera | Dugout canoe and outboard motor (pleasure boats) |
| Hunting rifles | Building/House |
| Undeveloped land |  |

Table SM7 – Classification of type dwelling categories into three ordinal levels (worst to best)

| **Characteristic** | **Category 1 (worst)** | **Category 2 (intermediate)** | **Category 3 (best)** |
| --- | --- | --- | --- |
| **Type of dwelling** | Hut / straw / mud brick / shack / tent / shed / tarpaulin | Detached / semi-detached house (solid construction) | Flat /apartment in a block of flats / building / multi-apartment |
|  | Traditional style / simple detached house | Terrace (compartmentalized) | Duplex / building (two-story house) |
|  | Room / servant quarters | Other | (other) Modern-style house |

Table SM8 – Classification of roof material categories into three ordinal levels (worst to best)

| **Characteristic** | **Category 1 (worst)** | **Category 2 (intermediate)** | **Category 3 (best)** |
| --- | --- | --- | --- |
| **Roof material** | Grass / leaves / bamboo / thatch / reed / mats / straw | Corrugated iron sheets (metal) / tin / zinc | Concrete (slab) / cement (slab) |
|  | Plastic canvas / sheeting | (sheet) Asbestos | Bricks |
|  | Mud and grass / wood |  | Tiles |
|  | Clay tiles / brick |  |  |
|  | Other |  |  |

Table SM9 – Classification of fuel for cooking categories into three ordinal levels (worst to best)

| **Characteristic** | **Category 1 (worst)** | **Category 2 (intermediate)** | **Category 3 (best)** |
| --- | --- | --- | --- |
| **Fuel for cooking** | Crop residue / leaves / dung / manure / sawdust / animal waste | Purchased firewood / wood | Biogas / butane / gas / LPG |
|  | Collected firewood / wood | Kerosene / Petroleum / Oil | Solar (energy) |
|  |  | Charcoal | Electricity |
|  |  | Paraffin |  |
|  |  | Other |  |
|  |  | No cooking / none |  |

Table SM10 – Classification of source of drinking water categories into four ordinal levels (worst to best)

| **Characteristic** | **Category 1 (worst)** | **Category 2** | **Category 3** | **Category 4 (best)** |
| --- | --- | --- | --- | --- |
| **Source of drinking water** | Surface water (dam, pond / lake, river / stream, canal, irrigation channels, reservoir) | Rainwater (collection) / gravity flow schemes | Tanker truck / bowser / cart with small tank / drum | Piped into yard / plot |
|  | Unprotected spring | Protected spring | Piped water to the neighbor | Piped into the dwelling |
|  | Open / unprotected well (public or in a yard / plot / concession area) | Protected well (in yard / plot or elsewhere) | Borehole (in yard / plot / Within the concession or elsewhere) | Bottled water / kiosk / retailer / sachet water |
|  | Street vendor | Communal standpipe / public taps | Borehole |  |
|  | Other |  |  |  |

Table SM11 – Classification of source of drinking water categories into three ordinal levels (worst to best)

| **Characteristic** | **Category 1 (worst)** | **Category 2 (intermediate)** | **Category 3 (best)** |
| --- | --- | --- | --- |
| **Source of drinking water** | Surface water (river, dam, lake, pond, stream, canal, irrigation channels) | Rainwater collection | Cart with small tank / drum |
|  | Unprotected dugwell | Tubewell / borehole | Bottled water |
|  | Unprotected spring | Protected dugwell | Tanker-truck |
|  | Other | Protected spring | Piped water |

Table SM12 – Classification of kitchen categories into three ordinal levels (worst to best)

| **Characteristic** | **Category 1 (worst)** | **Category 2 (intermediate)** | **Category 3 (best)** |
| --- | --- | --- | --- |
| **Kitchen** | A room used for traditional kitchen outside the housing unit | A room used for traditional kitchen inside housing unit | A room used for modern kitchen outside the housing unit |
|  | No kitchen | Other | A room used for modern kitchen inside housing unit |

Table SM13 – Classification of type of toilet categories into three ordinal levels (worst to best)

| **Characteristic** | **Category 1 (worst)** | **Category 2 (intermediate)** | **Category 3 (best)** |
| --- | --- | --- | --- |
| **Type of toilet** | No toilet (in nature) / no facility / bush / field | SANPLAT / ECOSAN latrines (paved, covered / uncovered) | (pour / outdoor / indoor / manual) Flush toilet / to pit latrine / to septic tank / to piped sewer system |
|  | Hanging toilet / hanging latrine | Simply / simple paved /slab latrines | VIP latrines (tiled / slab, ventilated) |
|  | Rudimentary pit / open hole | Pit latrine without slab / open pit | (washable) Pit latrine with slab |
|  | Public toilets | Other | Composting toilet |

Table SM14 – Classification of type of toilet categories into four ordinal levels (worst to best)

| **Characteristic** | **Category 1 (worst)** | **Category 2** | **Category 3** | **Category 4 (best)** |
| --- | --- | --- | --- | --- |
| **Type of toilet** | No facility / field / forest / bush | Hanging toilet / hanging latrine | Pit latrine with / without slab | Flush to dk where / to open drain / to piped sewer system / to pit latrine / to septic tank |
|  |  | Open pit | Container based sanitation | Ventilated improved pit latrine |
|  |  | Bucket | Composting toilet | Twin pit with slab |
|  |  | Other | Twin pit without slab |  |

Table SM15 – Classification of garbage collection categories into three ordinal levels (worst to best)

| **Characteristic** | **Category 1 (worst)** | **Category 2 (intermediate)** | **Category 3 (best)** |
| --- | --- | --- | --- |
| **Garbage collection** | Disposed of within household yard / plot / heaped in compound | Transported to the fields / disposed / heaped elsewhere | Collected by formal / informal (public) / service provider / gov / municipality / private company / rubbish bin |
|  | Use as fertilizer | Buried or burned elsewhere / in compound | Disposed of in designated waste disposable area |
|  | Illegal dump | Other | Public rubbish heap / dump |
|  | None |  | Rubbish pit / public trash can |

Table SM16 – Classification of walls material categories into three ordinal levels (worst to best)

| **Characteristic** |  | **Category 1 (worst)** | **Category 2 (intermediate)** | **Category 3 (best)** |
| --- | --- | --- | --- | --- |
| **Walls material** |  | Poles (bamboo), branches, grass, straw | Unburnt / burnt / baked / stabilized bricks with mud / cement / semi-hard adobe | (slab) Concrete and (blocks) cement and (masonry) stones |
|  |  | Compacted /rammed earth | Tin / iron / iron sheets / aluminum trays / sheets / sheets of metal | Cement blocks / concrete |
|  |  | Simple stones (traditional) | Recycled / reclaimed materials (planks, boards) | Concrete and stones |
|  |  | Mud and pole / stones | Wood |  |
|  |  | Mud, clod of earth | Other |  |
|  |  | No fence |  |  |
|  |  | Bench |  |  |

Table SM17 – Classification of walls material categories into four ordinal levels (worst to best)

| **Characteristic** | **Category 1 (worst)** | **Category 2** | **Category 3** | **Category 4 (best)** |
| --- | --- | --- | --- | --- |
| **Walls material** | Reed / bamboo | Wood and thatch | Corrugated iron sheet | Blocks, plastered with cement |
|  |  | Wood and mud | Asbestos | Blocks unplastered |
|  |  | Stone and mud | Bricks | Stone and cement |
|  |  | Chip wood | Steel |  |
|  |  | Wood |  |  |
|  |  | Stone |  |  |
|  |  | Other |  |  |

Table SM18 – Classification of wastewater disposal categories into three ordinal levels (worst to best)

| **Characteristic** | **Category 1 (worst)** | **Category 2 (intermediate)** | **Category 3 (best)** |
| --- | --- | --- | --- |
| **Wastewater disposal (sewage)** | In /on the street / (in) nature | Hole in the plot / ground | Retention tank / modern septic tank) |
|  |  | Other | Sewer pit (modern cesspool) / sewers |

Table SM19 – Classification of lighting categories into three ordinal levels (worst to best)

| **Characteristic** | **Category 1 (worst)** | **Category 2 (intermediate)** | **Category 3 (best)** |
| --- | --- | --- | --- |
| **Lighting** | (collected) Firewood / wood / boards | Battery / dry cell (torch) / mobile phone flashlight / ordinary torch | (electricity with) Solar panels |
|  | Torch / oil lamp / kerosene | Solar lamp / bulb / battery-powered/operated lamp | (from the grid) Electricity |
|  | Grass / crop residue | Private (electricity) generator | Gas (biogas) |
|  | Paraffin / candles | Purchased firewood |  |
|  | No lighting | Charcoal |  |
|  |  | Other |  |

Table SM20 – Classification of lighting categories into four ordinal levels (worst to best)

| **Characteristic** | **Category 1 (worst)** | **Category 2** | **Category 3** | **Category 4 (best)** |
| --- | --- | --- | --- | --- |
| **Lighting** | Candle / wax | Kerosene lamp (local kuraz) | Light from dry cell with switch / electrical battery | Electricity meter – private |
|  | Firewood | Other | Kerosene light (imported) | Solar energy |
|  |  |  | Electricity from generator |  |
|  |  |  | Electricity meter – shared |  |
|  |  |  | Lantern |  |
|  |  |  | Biogas |  |

Table SM21 – Classification of handwashing place categories into three ordinal levels (worst to best)

| **Characteristic** | **Category 1 (worst)** | **Category 2 (intermediate)** | **Category 3 (best)** |
| --- | --- | --- | --- |
| **Handwashing place** | Not observed | Observed, not fixed | Observed, fixed location |

Figure SM1. Percentage distributions of urban, rural, main urban center, and 100% rural district households by quintile for each index – Benin

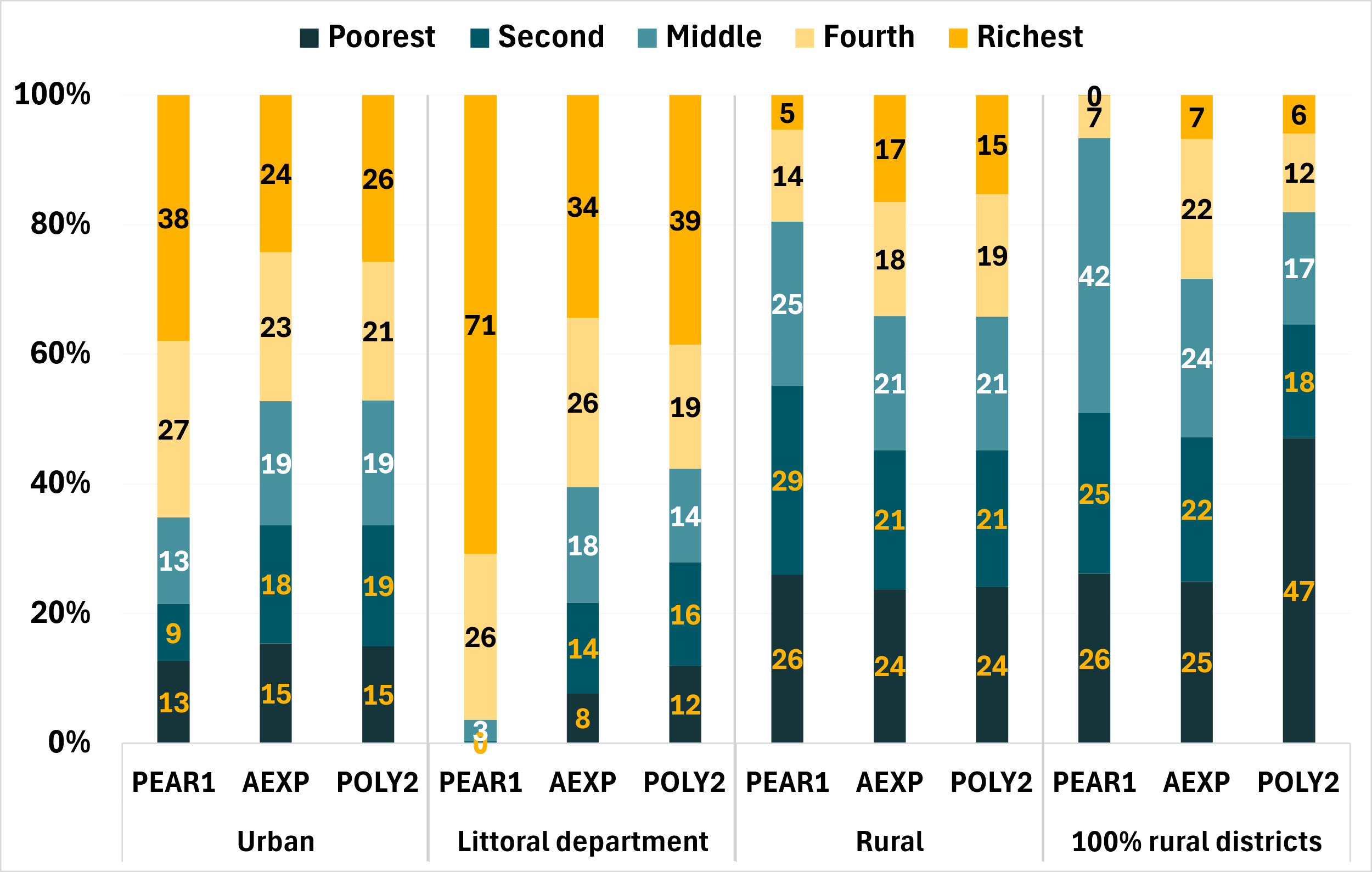

Figure SM2. Percentage distributions of urban, rural, main urban center, and 100% rural district households by quintile for each index – Burkina Faso

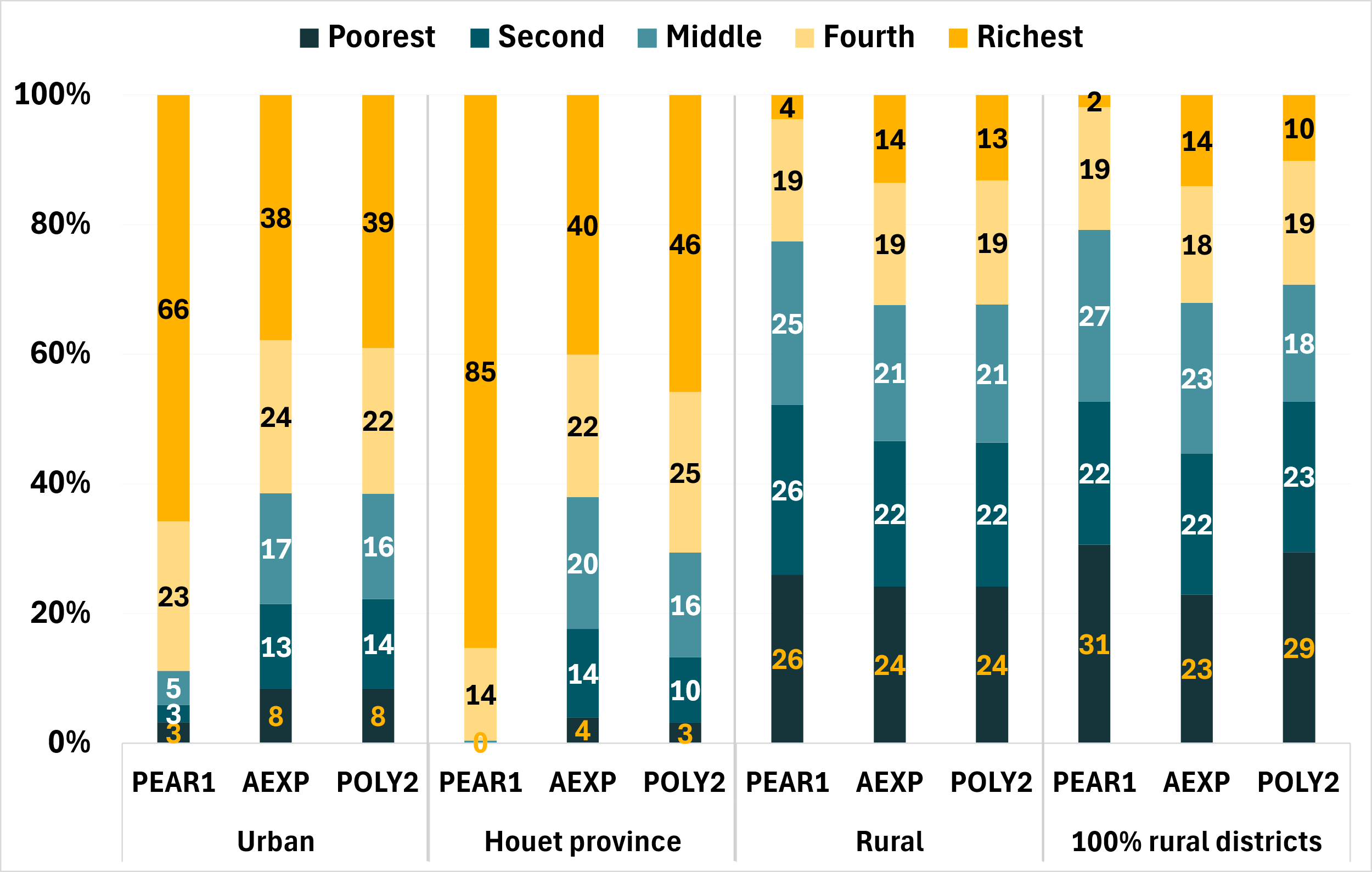

Figure SM3. Percentage distributions of urban, rural, main urban center, and 100% rural district households by quintile for each index – Côte d’Ivoire

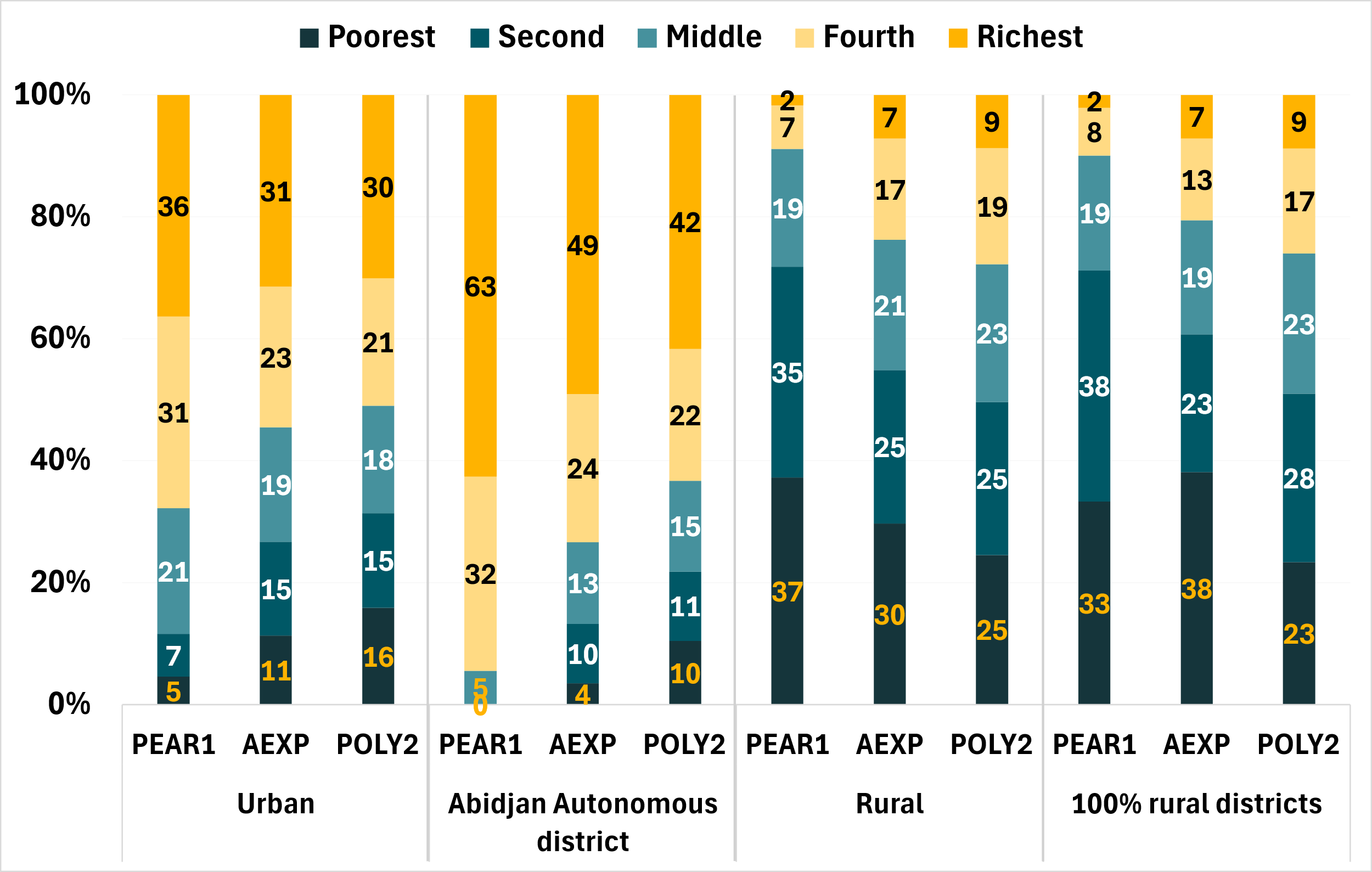

Figure SM4. Percentage distributions of urban, rural, main urban center, and 100% rural district households by quintile for each index – Ethiopia

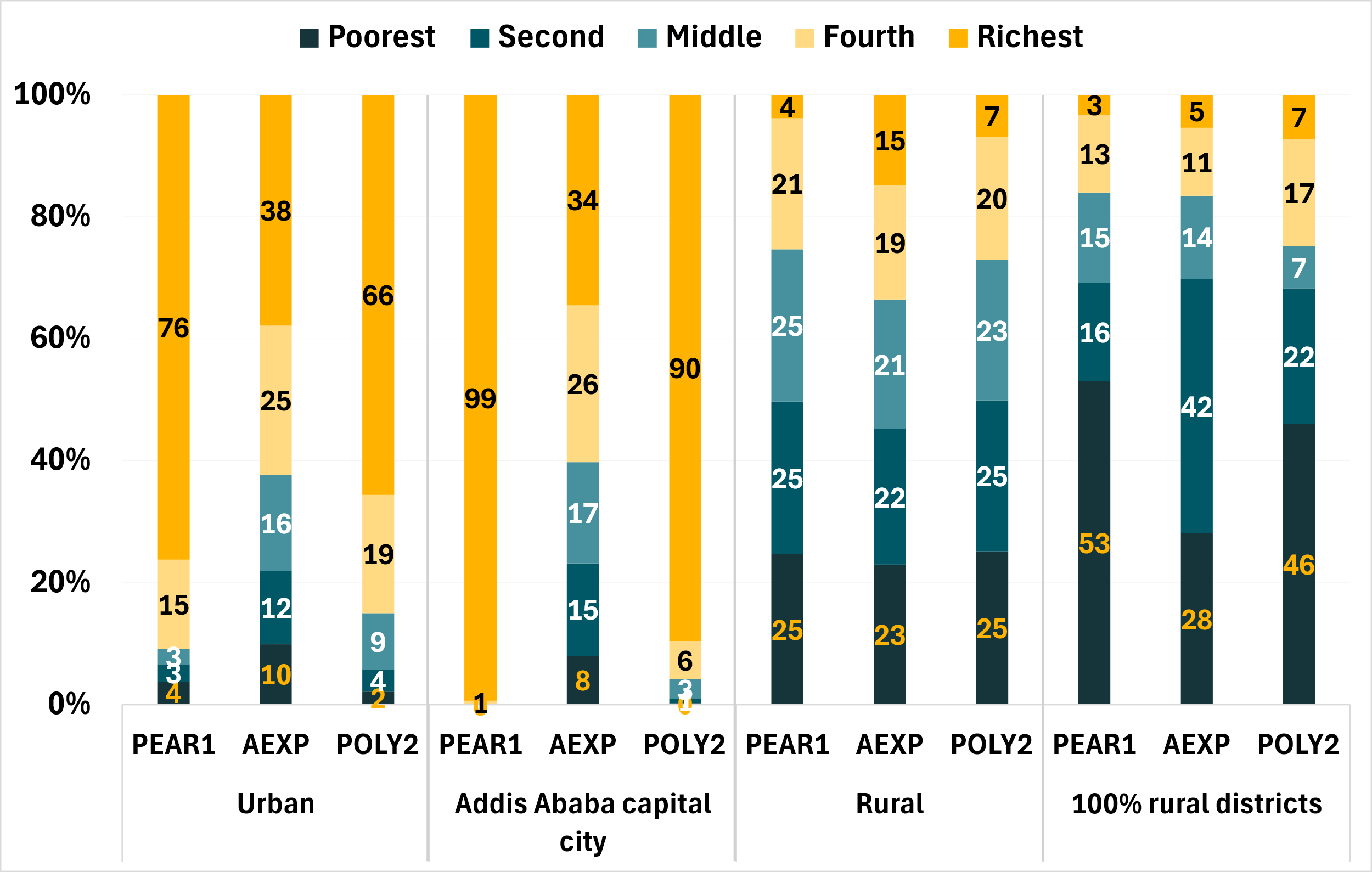

Figure SM5. Percentage distributions of urban, rural, main urban center, and 100% rural district households by quintile for each index – Guinea-Bissau

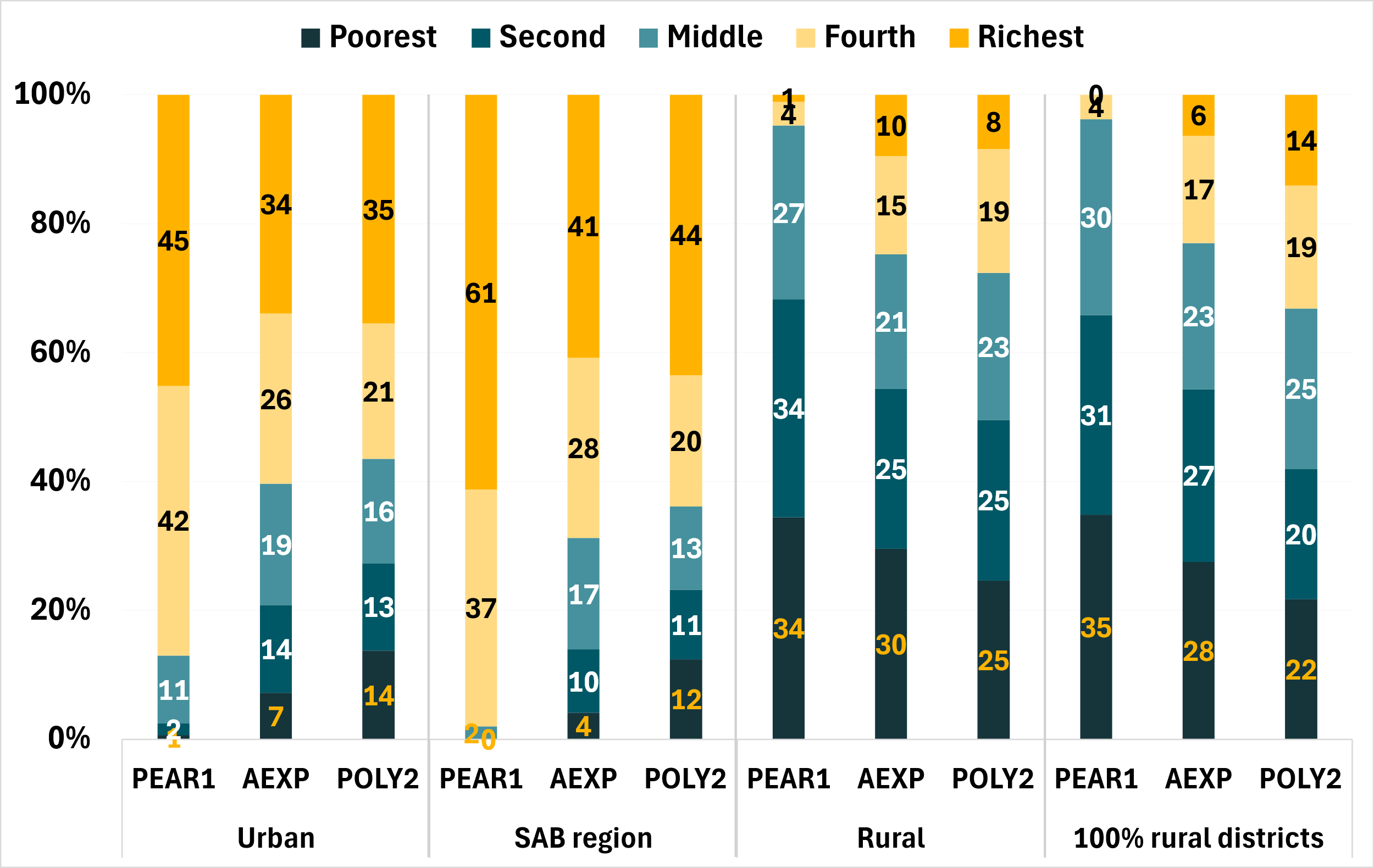

Figure SM6. Percentage distributions of urban, rural, main urban center, and 100% rural district households by quintile for each index – Malawi

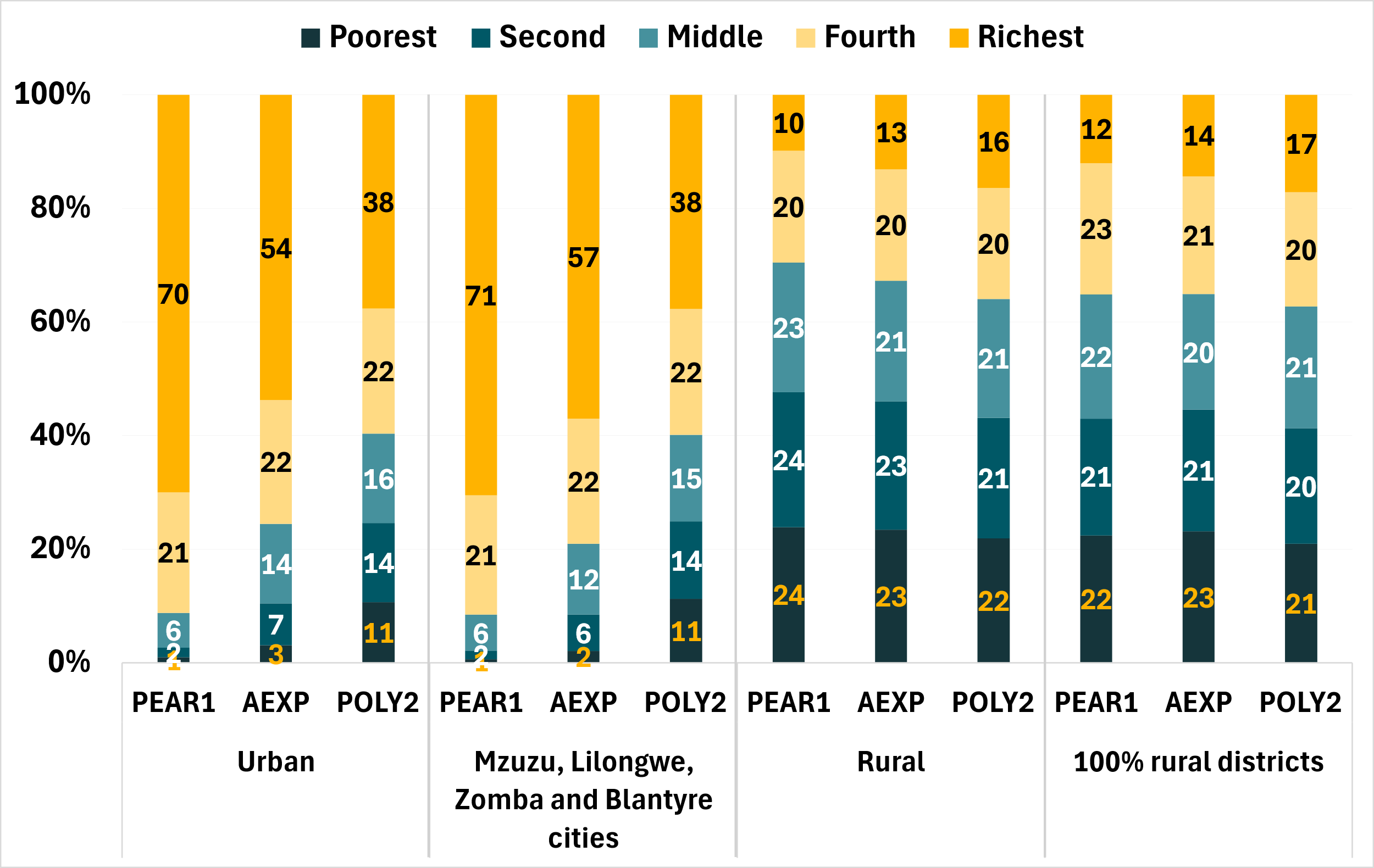

Figure SM7. Percentage distributions of urban, rural, main urban center, and 100% rural district households by quintile for each index – Mali

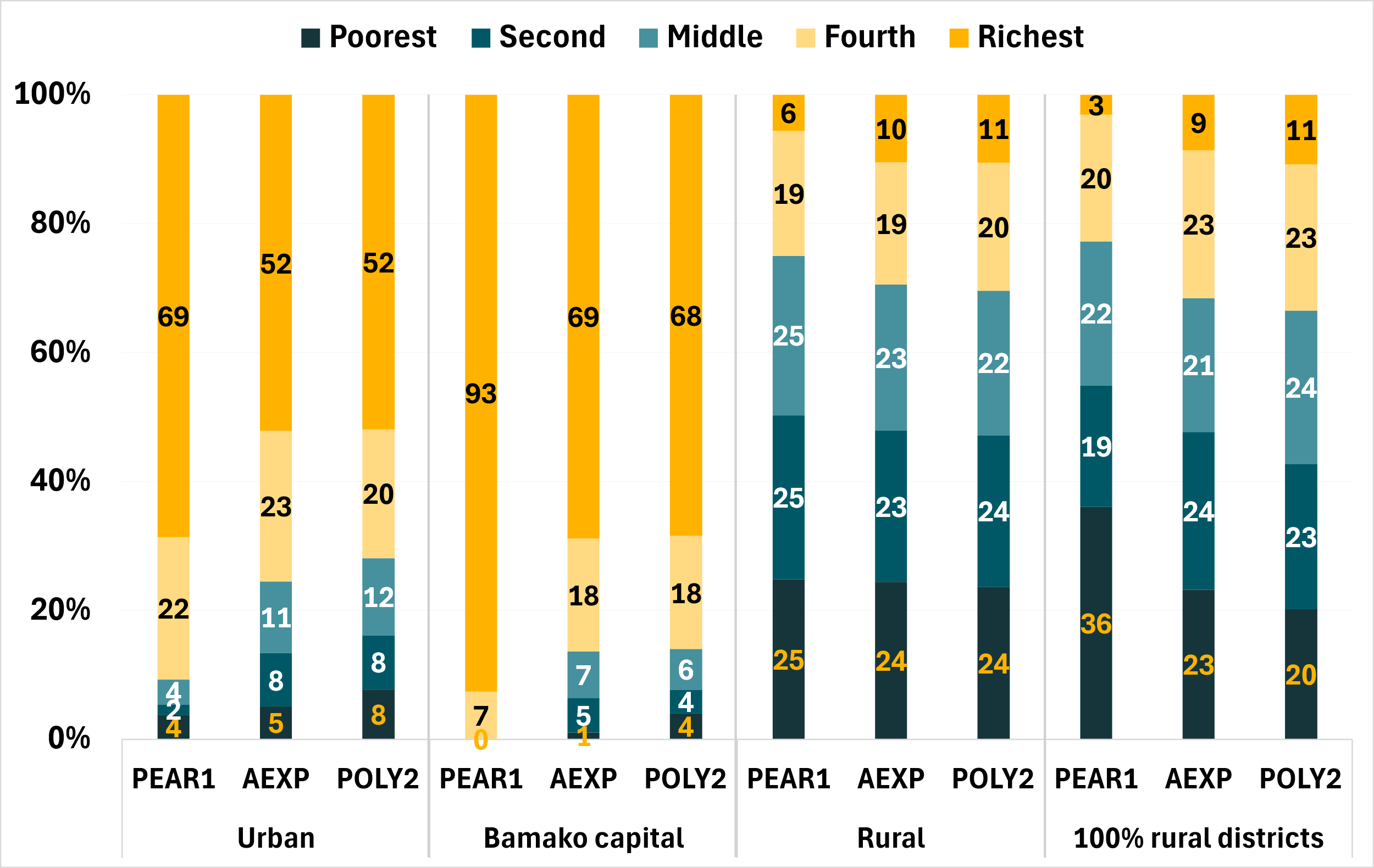

Figure SM8. Percentage distributions of urban, rural, main urban center, and 100% rural district households by quintile for each index – Niger

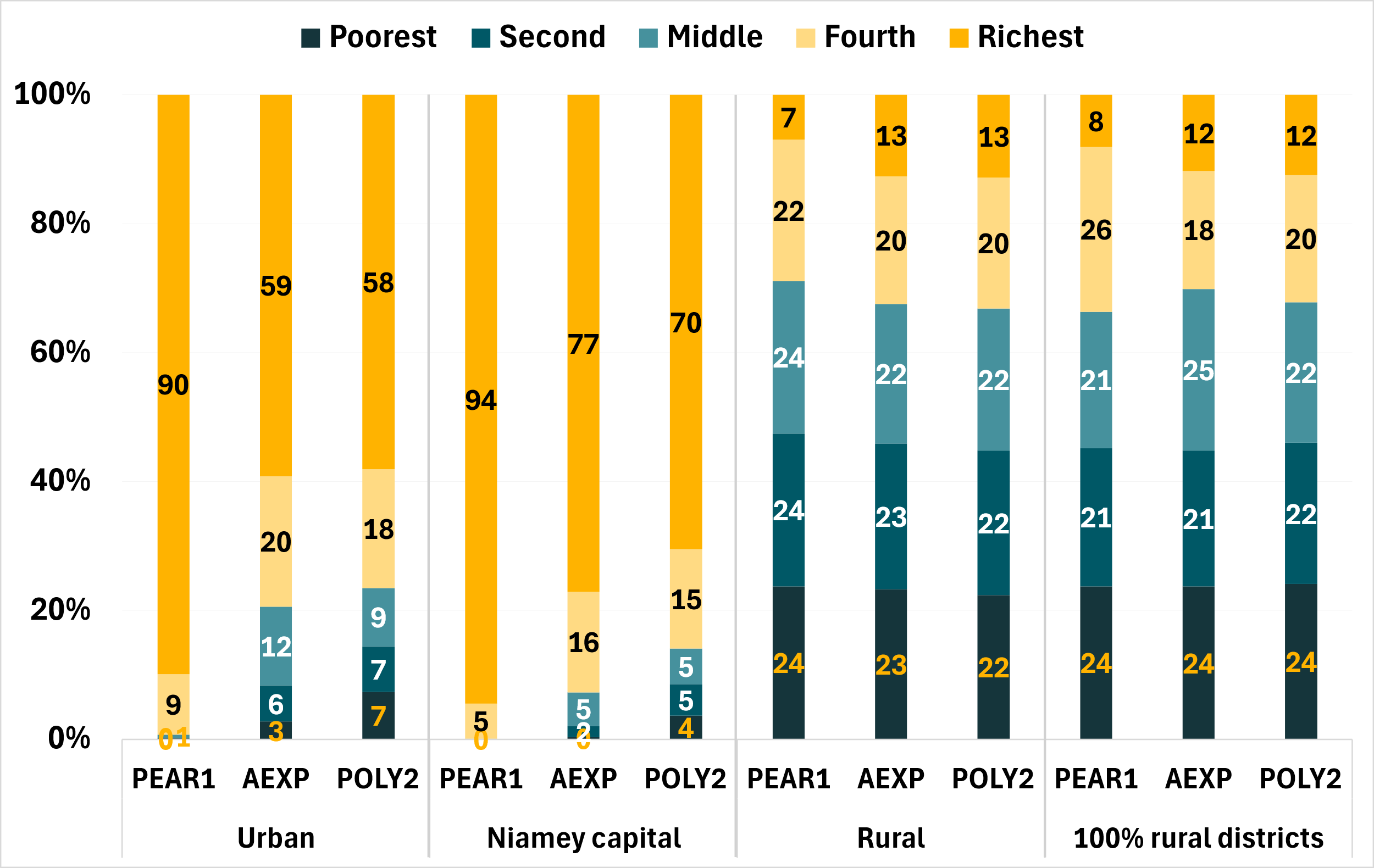

Figure SM9. Percentage distributions of urban, rural, main urban center, and 100% rural district households by quintile for each index – Senegal

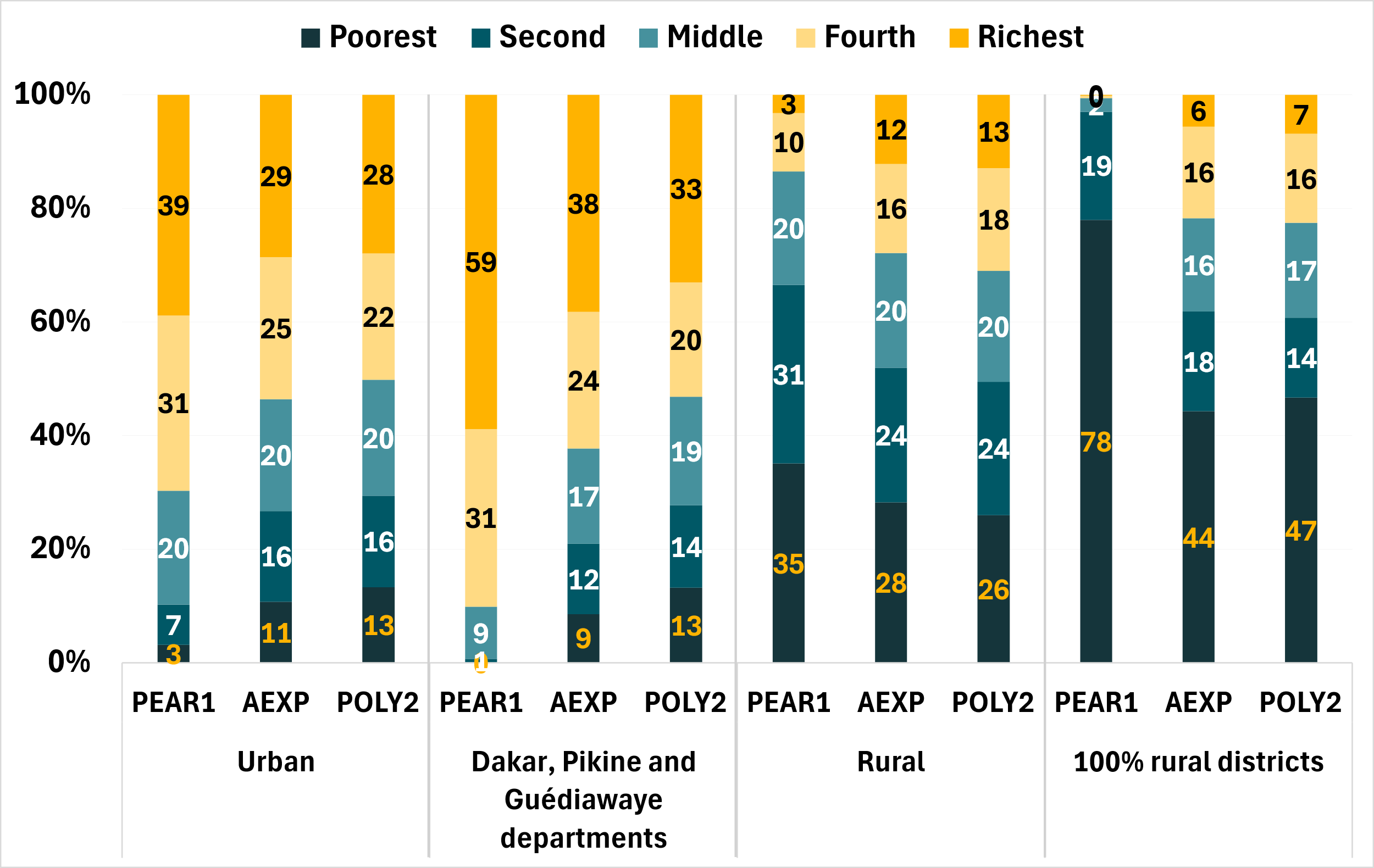

Figure SM10. Percentage distributions of urban, rural, main urban center, and 100% rural district households by quintile for each index – Togo

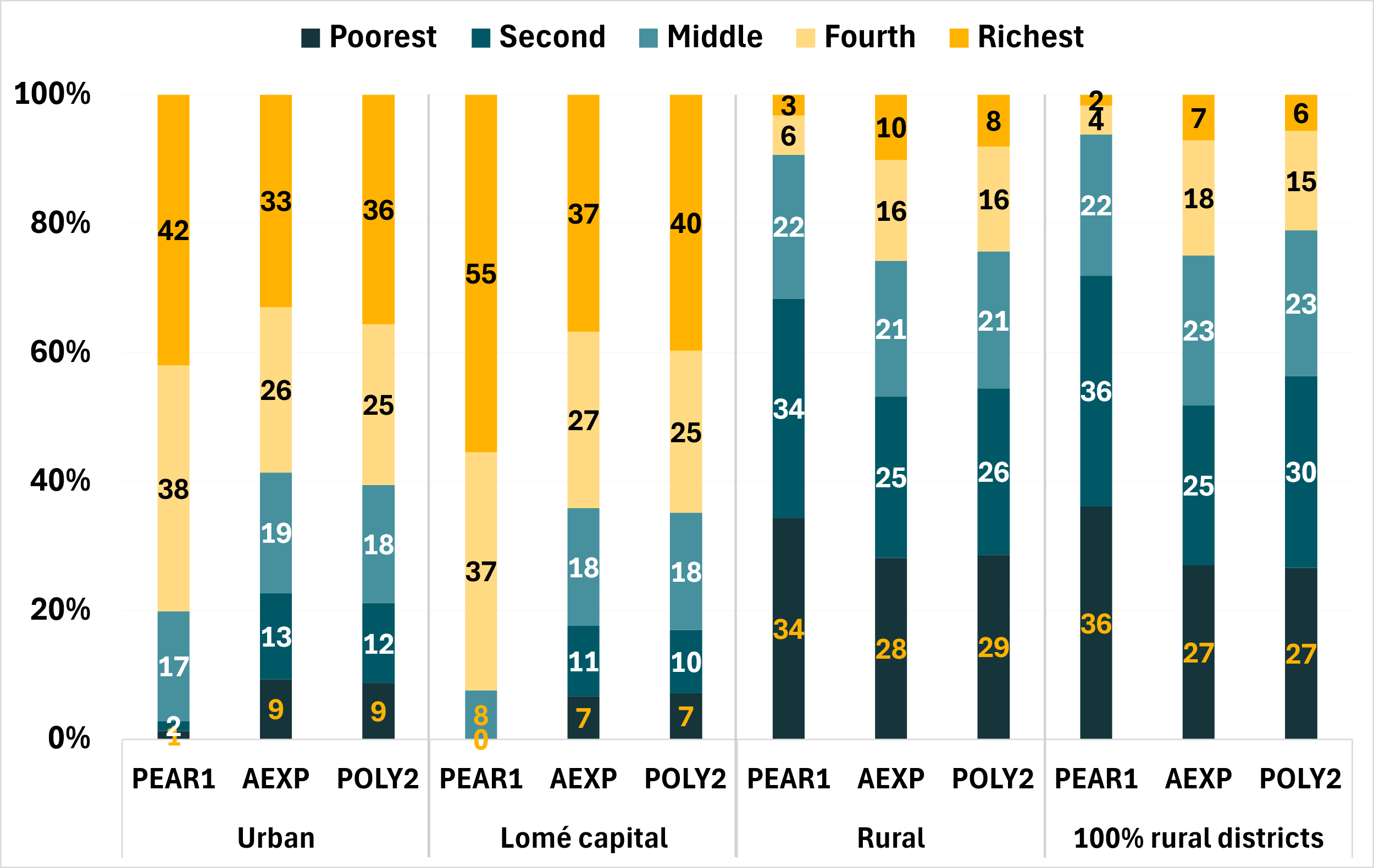

Figure SM11. Percentage distributions of urban, rural, main urban center, and 100% rural district households by quintile for each index – Uganda

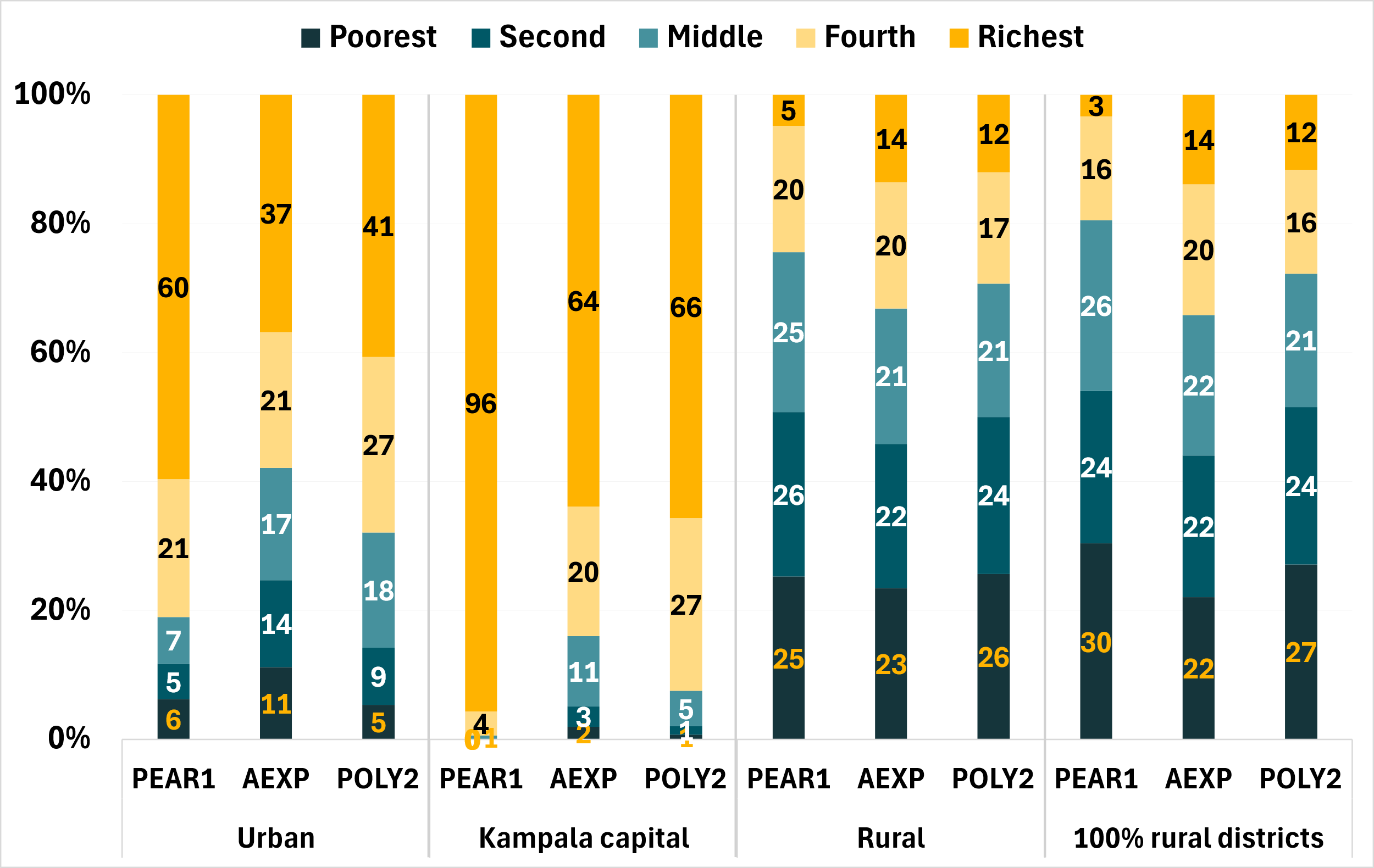

Table SM22. Comparison of approaches regarding observed agreement and the Kappa coefficient at the national level – with confidence intervals

|  |  | **National** | | | | | | |
| --- | --- | --- | --- | --- | --- | --- | --- | --- |
|  |  | **Agreement** | | |  | **Kappa** | | |
| **Survey** |  | **AEXP x PEAR1 (95% CI)** | **AEXP x POLY2 (95% CI)** | **PEAR1 x POLY2** |  | **AEXP x PEAR1 (95% CI)** | **AEXP x POLY2 (95% CI)** | **PEAR1 x POLY2** |
| Benin (2021) |  | 70.8% (69.2%, 72.3%) | 74.8% (73.3%, 76.3%) | 73.1% |  | 26.9% (25.4%, 28.4%) | 37.0% (35.5%, 38.5%) | 32.7% |
| Burkina Faso (2021) |  | 74.0% (72.4%, 75.6%) | 79.8% (78.2%, 81.4%) | 79.4% |  | 35.1% (33.5%, 36.7%) | 49.5% (47.9%, 51.1%) | 48.5% |
| Côte d’Ivoire (2021) |  | 72.3% (71.2%, 73.5%) | 76.1% (74.9%, 77.3%) | 76.2% |  | 30.9% (29.7%, 32.1%) | 40.3% (39.1%, 41.5%) | 40.6% |
| Ethiopia (2021) |  | 69.4% (67.5%, 71.3%) | 72.2% (70.3%, 74.1%) | 88.6% |  | 23.5% (21.5%, 25.4%) | 30.6% (28.6%, 32.5%) | 71.6% |
| Guinea-Bissau (2021) |  | 75.2% (73.4%, 77.1%) | 77.9% (76.1%, 79.8%) | 76.2% |  | 38.1% (36.2%, 39.9%) | 44.9% (43.0%, 46.7%) | 40.4% |
| Mali (2021) |  | 77.0% (75.3%, 78.8%) | 78.5% (76.8%, 80.3%) | 79.7% |  | 42.6% (40.9%, 44.4%) | 46.3% (44.5%, 48.0%) | 49.3% |
| Malawi (2019) |  | 78.6% (77.2%, 79.9%) | 77.8% (76.5%, 79.2%) | 83.7% |  | 46.5% (45.1%, 47.8%) | 44.6% (43.2%, 45.9%) | 59.2% |
| Niger (2021) |  | 78.4% (76.7%, 80.0%) | 78.4% (76.8%, 80.1%) | 77.8% |  | 45.9% (44.3%, 47.6%) | 46.1% (44.4%, 47.7%) | 44.4% |
| Senegal (2021) |  | 77.2% (75.6%, 78.8%) | 80.9% (79.3%, 82.5%) | 80.4% |  | 43.0% (41.4%, 44.6%) | 52.3% (50.7%, 53.9%) | 51.1% |
| Togo (2021) |  | 72.1% (70.4%, 73.8%) | 76.3% (74.6%, 78.0%) | 77.3% |  | 30.3% (28.6%, 32.0%) | 40.8% (39.1%, 42.5%) | 43.3% |
| Tanzania (2021) |  | 71.9% (70.0%, 73.9%) | 79.2% (77.2%, 81.1%) | 80.5% |  | 29.8% (27.9%, 31.8%) | 47.9% (45.9%, 49.9%) | 51.3% |
| Uganda (2019) |  | 71.4% (68.8%, 73.9%) | 76.0% (73.4%, 78.6%) | 83.2% |  | 28.4% (25.8%, 30.9%) | 40.0% (37.4%, 42.6%) | 57.9% |
| **Average** |  | **74.0%** | **77.3%** |  |  | **35.1%** | **43.3%** |  |

Table SM23. Comparison of approaches regarding observed agreement and the Kappa coefficient at the urban level – with confidence intervals

|  |  | **Urban** | | | | | | |
| --- | --- | --- | --- | --- | --- | --- | --- | --- |
|  |  | **Agreement** | | |  | **Kappa** | | |
| **Survey** |  | **AEXP x PEAR1 (95% CI)** | **AEXP x POLY2 (95% CI)** | **PEAR1 x POLY2** |  | **AEXP x PEAR1 (95% CI)** | **AEXP x POLY2 (95% CI)** | **PEAR1 x POLY2** |
| Benin (2021) |  | 71.2% (69.1%, 73.3%) | 76.1% (73.9%, 78.3%) | 74.5% |  | 26.9% (24.8%, 29.0%) | 40.0% (37.8%, 42.1%) | 35.7% |
| Burkina Faso (2021) |  | 74.2% (72.1%, 76.4%) | 80.5% (78.1%, 82.8%) | 80.5% |  | 29.2% (27.1%, 31.4%) | 49.6% (47.2%, 52.0%) | 46.7% |
| Côte d’Ivoire (2021) |  | 73.8% (72.1%, 75.5%) | 77.6% (75.7%, 79.5%) | 74.5% |  | 28.7% (27.0%, 30.5%) | 44.3% (42.4%, 46.2%) | 33.8% |
| Ethiopia (2021) |  | 68.5% (66.2%, 70.7%) | 72.9% (70.5%, 75.3%) | 86.9% |  | 11.6% (9.33%, 13.8%) | 25.1% (22.7%, 27.5%) | 55.2% |
| Guinea-Bissau (2021) |  | 78.2% (75.8%, 80.6%) | 79.7% (76.7%, 82.8%) | 76.4% |  | 26.9% (24.4%, 29.3%) | 45.3% (42.2%, 48.3%) | 29.5% |
| Mali (2021) |  | 78.6% (76.1%, 81.1%) | 81.5% (78.9%, 84.2%) | 82.7% |  | 38.2% (35.7%, 40.7%) | 50.9% (48.2%, 53.5%) | 51.7% |
| Malawi (2019) |  | 83.8% (80.9%, 86.6%) | 80.5% (77.6%, 83.4%) | 76.6% |  | 33.9% (31.1%, 36.7%) | 43.8% (40.9%, 46.7%) | 29.9% |
| Niger (2021) |  | 82.9% (80.5%, 85.3%) | 81.8% (79.1%, 84.5%) | 82.8% |  | 32.3% (30.0%, 34.7%) | 44.9% (42.2%, 47.6%) | 40.3% |
| Senegal (2021) |  | 77.4% (75.4%, 79.5%) | 81.2% (79.1%, 83.4%) | 80.7% |  | 38.1% (36.1%, 40.2%) | 51.6% (49.5%, 53.8%) | 47.7% |
| Togo (2021) |  | 74.3% (72.1%, 76.4%) | 77.6% (74.8%, 80.3%) | 81.0% |  | 22.8% (20.6%, 24.9%) | 39.2% (36.5%, 42.0%) | 39.0% |
| Tanzania (2021) |  | 74.1% (71.5%, 76.6%) | 81.1% (78.3%, 84.0%) | 82.4% |  | 21.6% (19.1%, 24.2%) | 45.8% (43.0%, 48.7%) | 40.6% |
| Uganda (2019) |  | 70.6% (66.1%, 75.1%) | 78.4% (73.4%, 83.4%) | 82.3% |  | 20.1% (15.6%, 24.6%) | 41.0% (36.0%, 46.0%) | 41.8% |
| **Average** |  | **75.6%** | **79.1%** |  |  | **27.5%** | **43.5%** |  |

Table SM24. Comparison of approaches regarding observed agreement and the Kappa coefficient at the urban level – with confidence intervals

|  |  | **Rural** | | | | | | |
| --- | --- | --- | --- | --- | --- | --- | --- | --- |
|  |  | **Agreement** | | |  | **Kappa** | | |
| **Survey** |  | **AEXP x PEAR1 (95% CI)** | **AEXP x POLY2 (95% CI)** | **PEAR1 x POLY2** |  | **AEXP x PEAR1 (95% CI)** | **AEXP x POLY2 (95% CI)** | **PEAR1 x POLY2** |
| Benin (2021) |  | 70.3% (68.3%, 72.3%) | 73.5% (71.4%, 75.6%) | 71.7% |  | 18.6% (16.7%, 20.6%) | 31.4% (29.3%, 33.5%) | 21.5% |
| Burkina Faso (2021) |  | 73.9% (71.9%, 75.9%) | 79.2% (77.0%, 81.3%) | 78.4% |  | 26.0% (24.0%, 27.9%) | 44.6% (42.5%, 46.8%) | 37.5% |
| Côte d’Ivoire (2021) |  | 71.3% (69.9%, 72.8%) | 75.1% (73.6%, 76.7%) | 77.5% |  | 19.5% (18.1%, 21.0%) | 34.2% (32.7%, 35.7%) | 37.7% |
| Ethiopia (2021) |  | 70.5% (68.2%, 72.7%) | 71.5% (69.0%, 74.0%) | 90.7% |  | 14.0% (11.8%, 16.2%) | 19.2% (16.7%, 21.7%) | 63.8% |
| Guinea-Bissau (2021) |  | 73.5% (71.3%, 75.6%) | 76.9% (74.6%, 79.2%) | 76.0% |  | 19.7% (17.6%, 21.8%) | 36.6% (34.3%, 38.9%) | 29.3% |
| Mali (2021) |  | 75.8% (73.6%, 78.0%) | 76.0% (73.6%, 78.3%) | 77.2% |  | 23.4% (21.2%, 25.6%) | 30.8% (28.5%, 33.1%) | 29.4% |
| Malawi (2019) |  | 77.3% (75.8%, 78.8%) | 77.2% (75.7%, 78.6%) | 85.4% |  | 38.4% (37.0%, 39.9%) | 40.9% (39.4%, 42.4%) | 61.5% |
| Niger (2021) |  | 75.6% (73.5%, 77.6%) | 76.3% (74.3%, 78.4%) | 74.7% |  | 21.3% (19.2%, 23.3%) | 30.5% (28.4%, 32.5%) | 19.7% |
| Senegal (2021) |  | 76.9% (74.7%, 79.1%) | 80.6% (78.2%, 83.0%) | 80.1% |  | 35.8% (33.6%, 38.0%) | 49.0% (46.6%, 51.3%) | 45.2% |
| Togo (2021) |  | 70.8% (68.8%, 72.7%) | 75.5% (73.4%, 77.7%) | 75.0% |  | 16.7% (14.8%, 18.7%) | 34.1% (32.0%, 36.2%) | 26.3% |
| Tanzania (2021) |  | 70.1% (67.7%, 72.5%) | 77.5% (74.9%, 80.1%) | 78.9% |  | 12.5% (10.1%, 14.9%) | 37.8% (35.2%, 40.5%) | 33.3% |
| Uganda (2019) |  | 71.6% (68.8%, 74.4%) | 75.2% (72.3%, 78.1%) | 83.5% |  | 22.8% (19.9%, 25.6%) | 34.9% (32.0%, 37.8%) | 53.6% |
| **Average** |  | **73.1%** | **76.2%** |  |  | **22.4%** | **35.3%** |  |

Figure SM12. Proportion of households with at least one member with secondary or higher education by wealth quintile, index, and geographic area – Benin

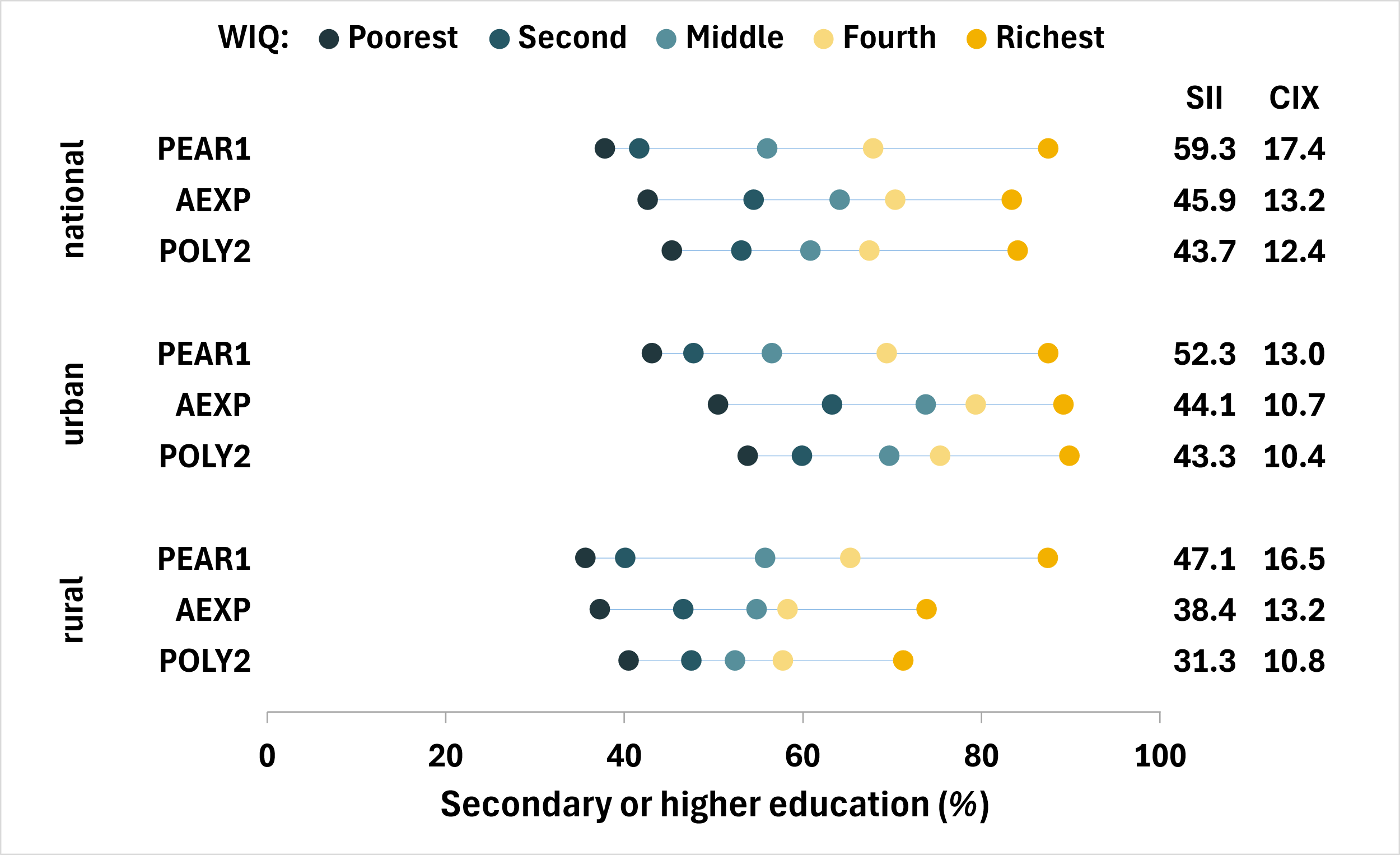

Figure SM13. Proportion of households with at least one member with secondary or higher education by wealth quintile, index, and geographic area – Burkina Faso

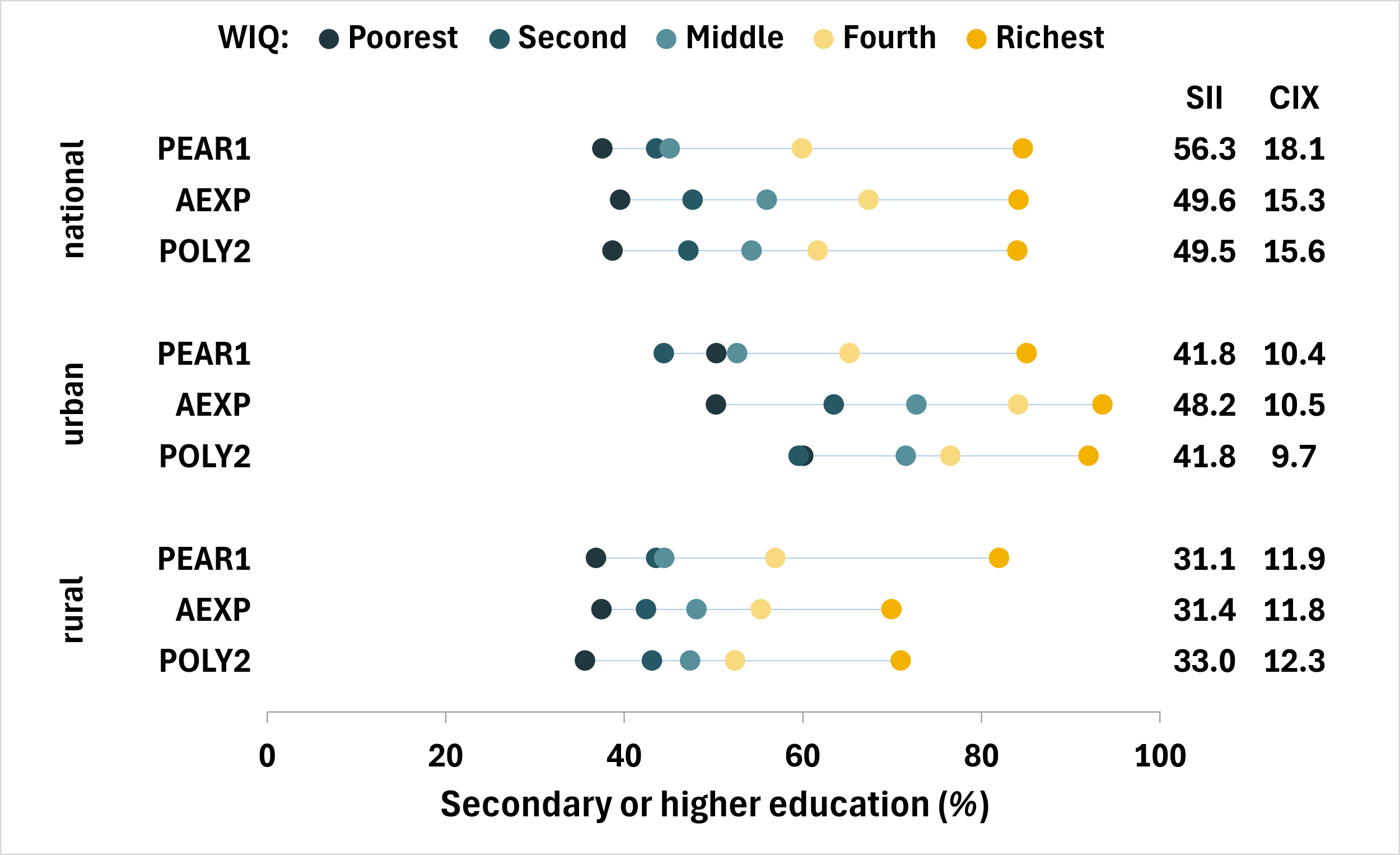

Figure SM14. Proportion of households with at least one member with secondary or higher education by wealth quintile, index, and geographic area – Ethiopia

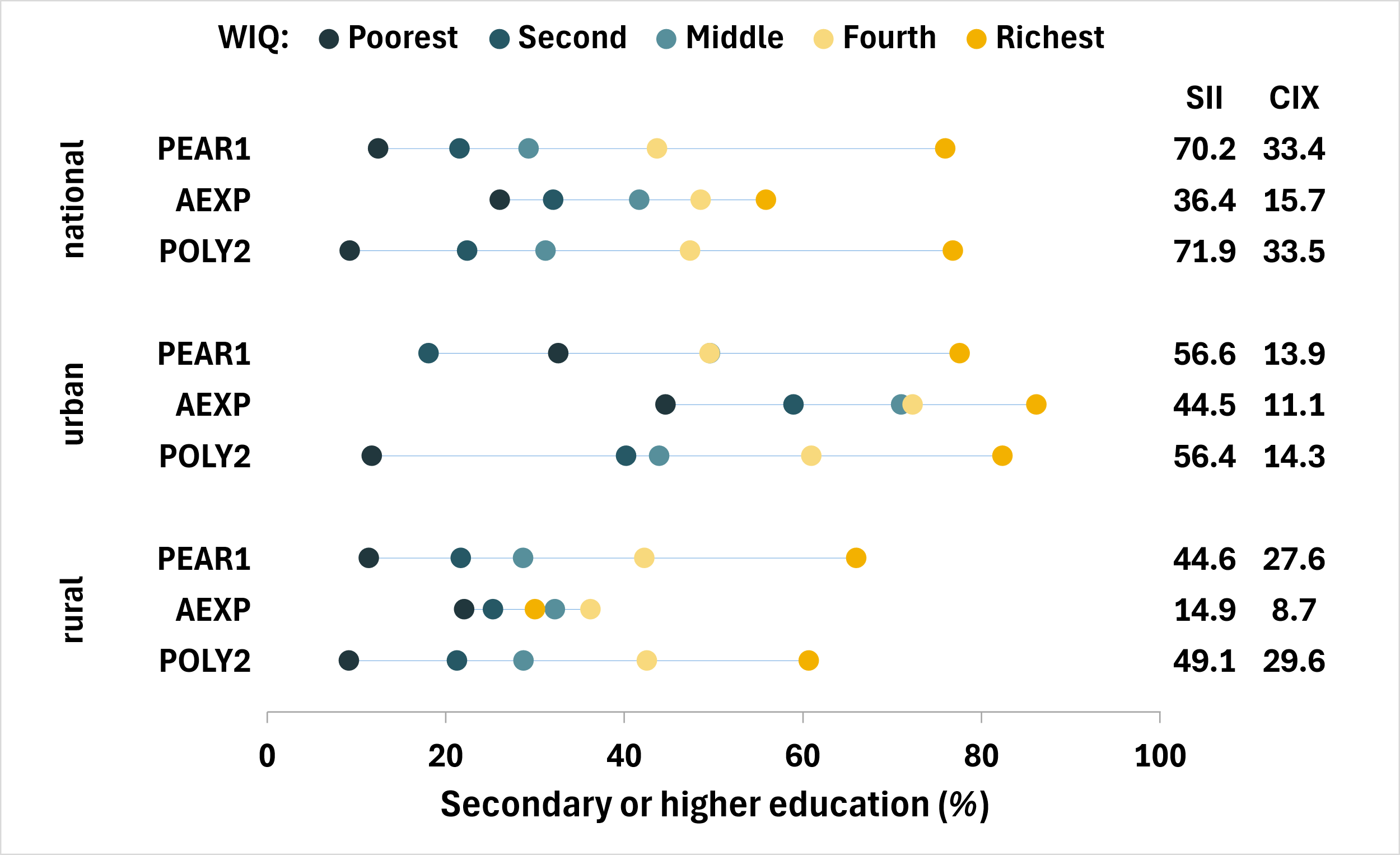

Figure SM15. Proportion of households with at least one member with secondary or higher education by wealth quintile, index, and geographic area – Guinea-Bissau

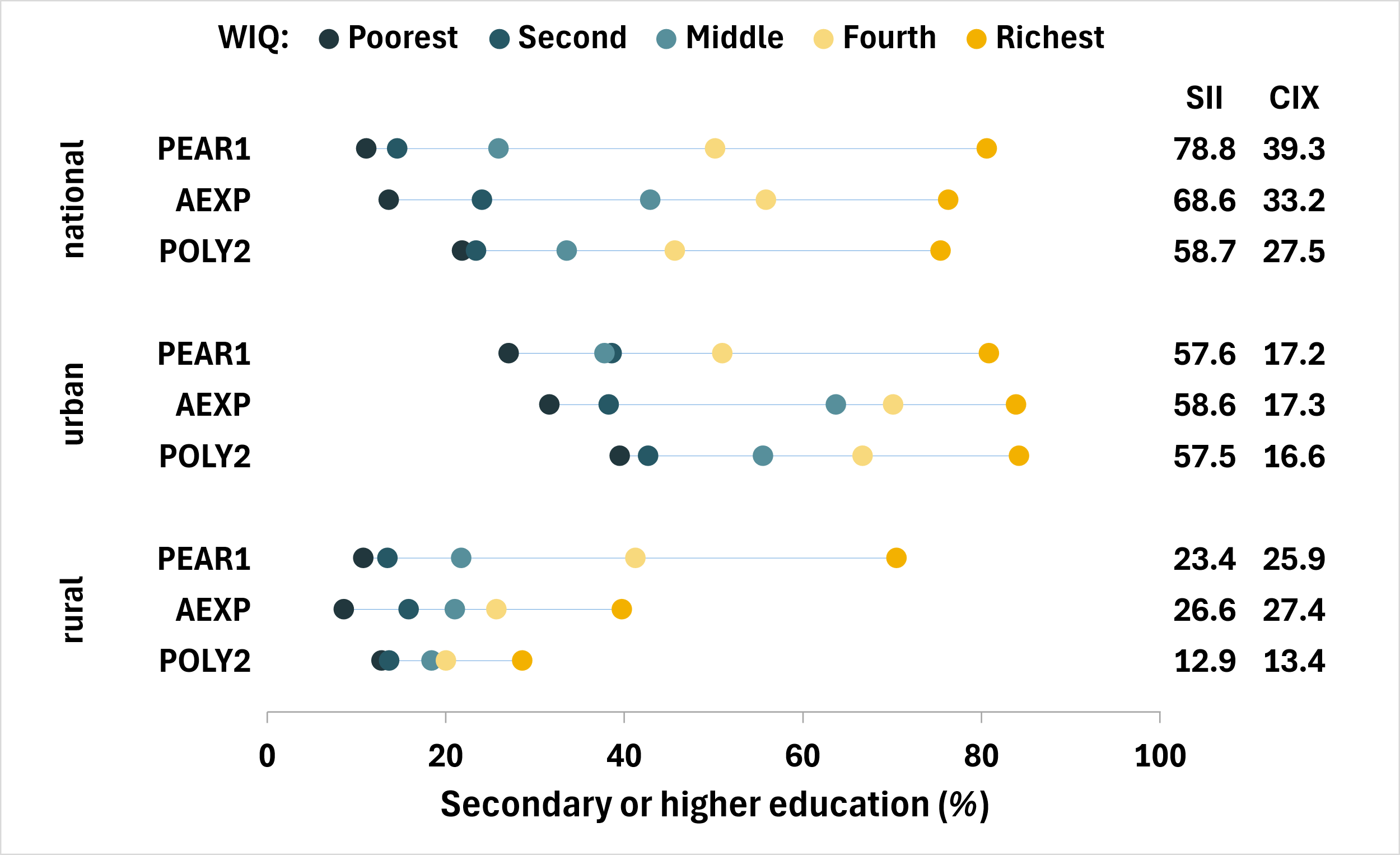

Figure SM16. Proportion of households with at least one member with secondary or higher education by wealth quintile, index, and geographic area – Malawi

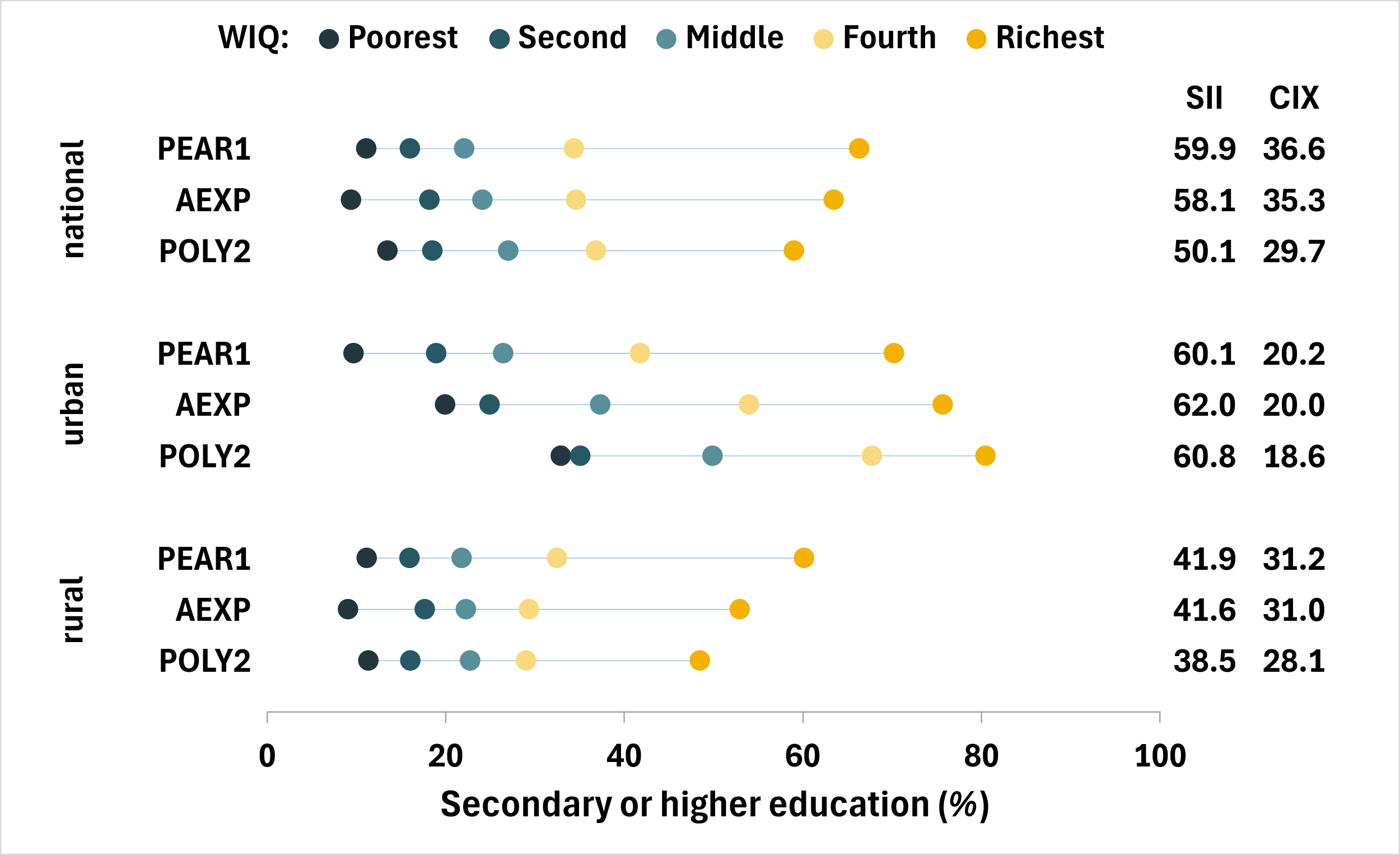

Figure SM17. Proportion of households with at least one member with secondary or higher education by wealth quintile, index, and geographic area – Mali

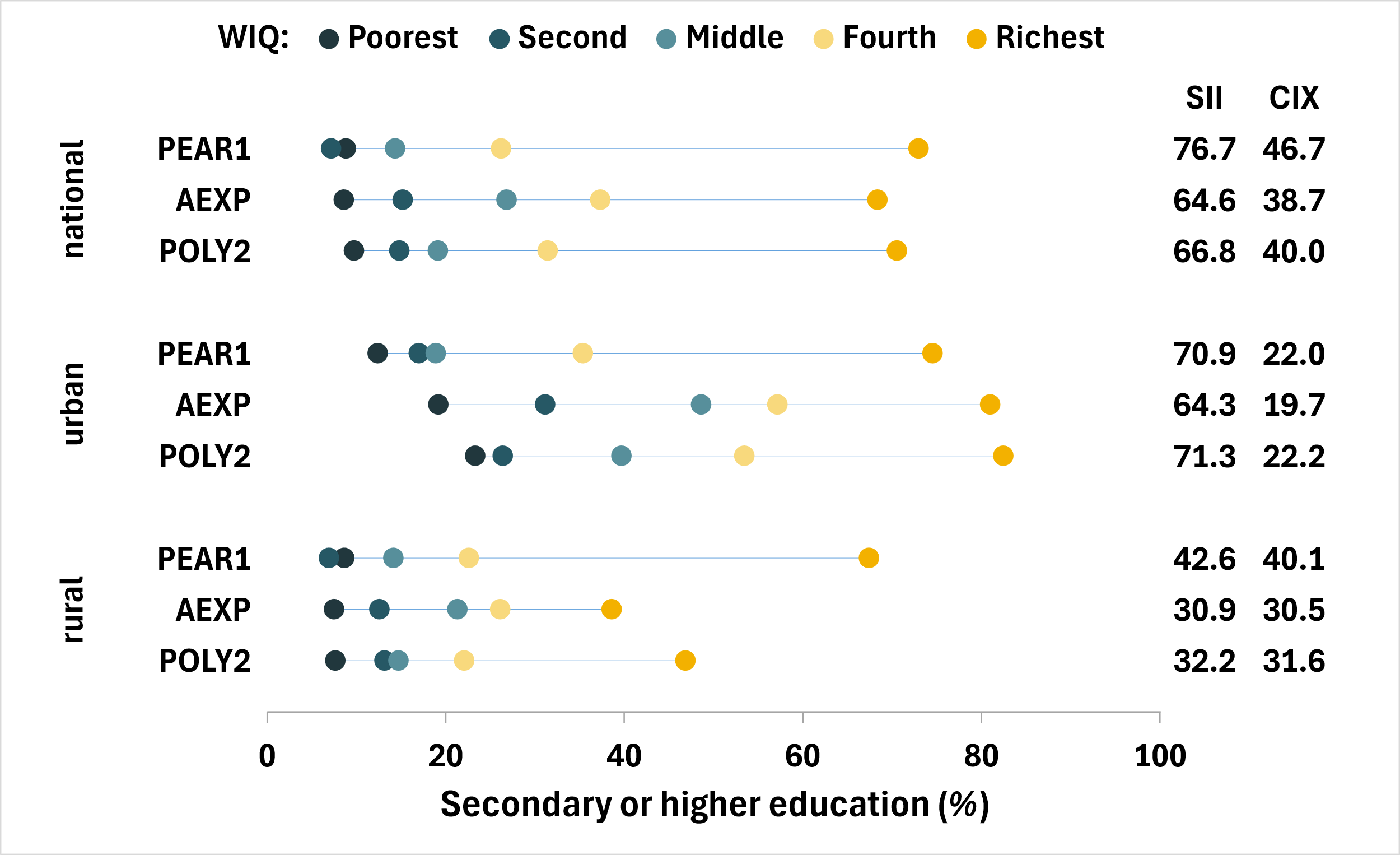

Figure SM18. Proportion of households with at least one member with secondary or higher education by wealth quintile, index, and geographic area – Niger

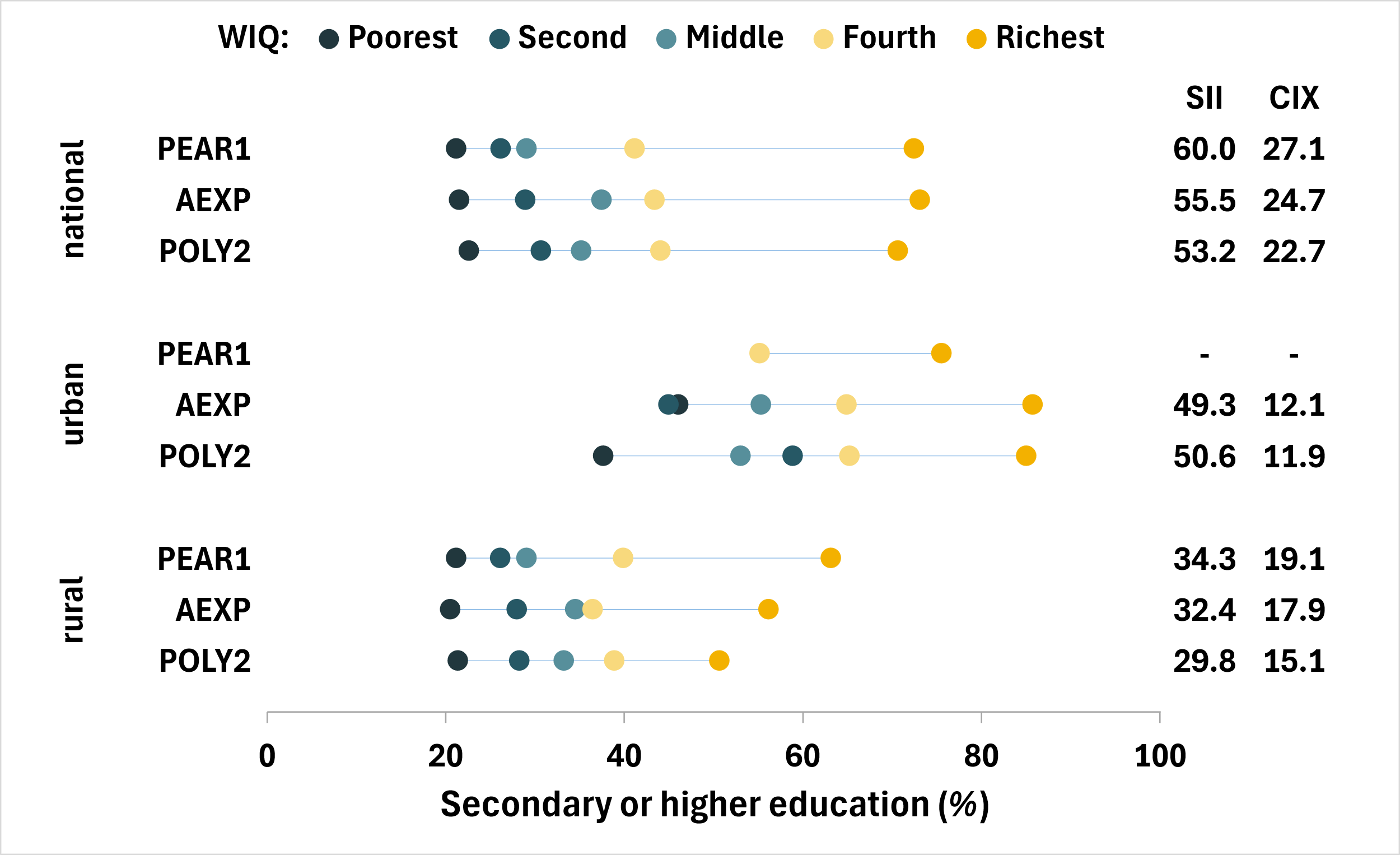

Figure SM19. Proportion of households with at least one member with secondary or higher education by wealth quintile, index, and geographic area – Senegal

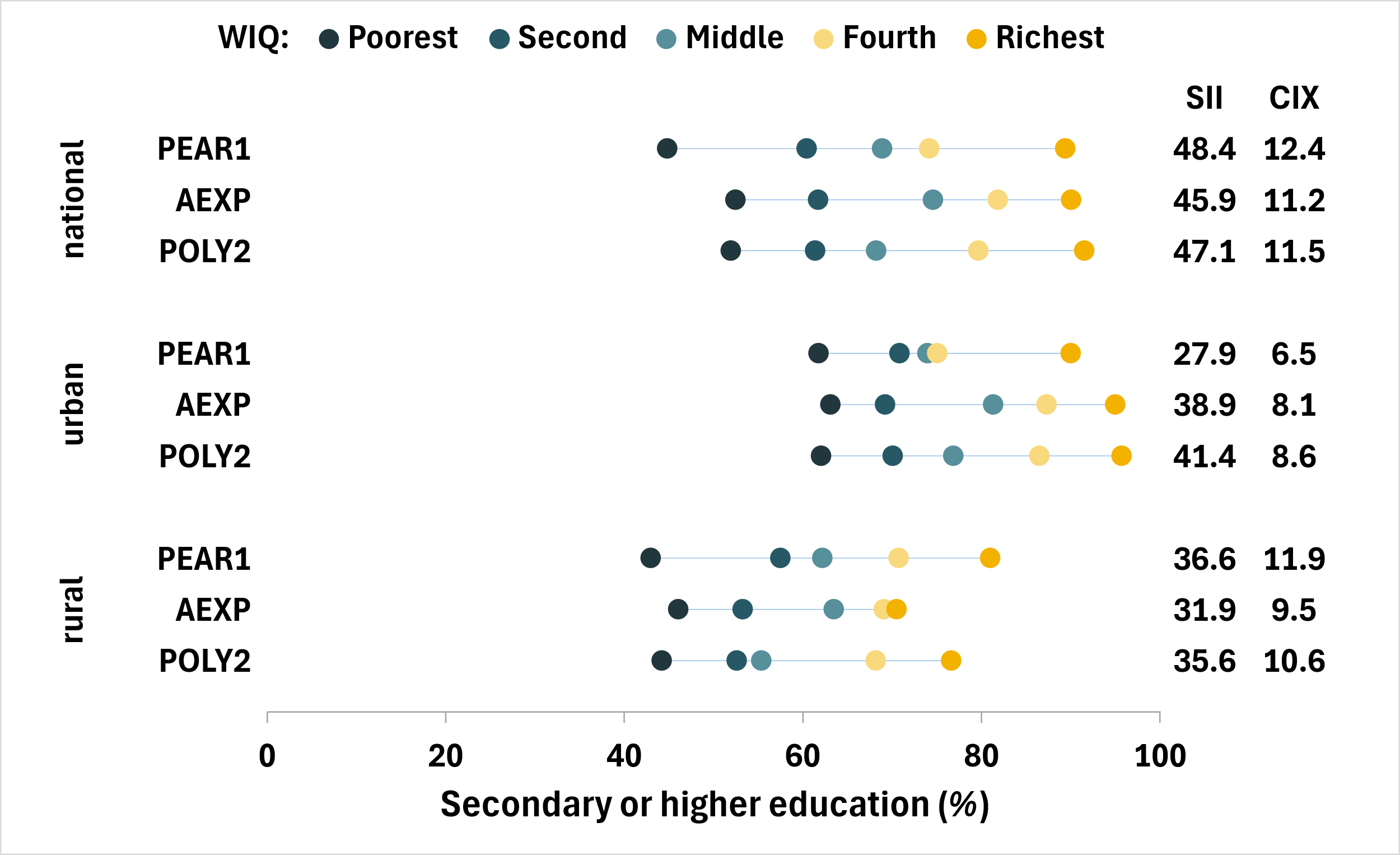

Figure SM20. Proportion of households with at least one member with secondary or higher education by wealth quintile, index, and geographic area – Tanzania

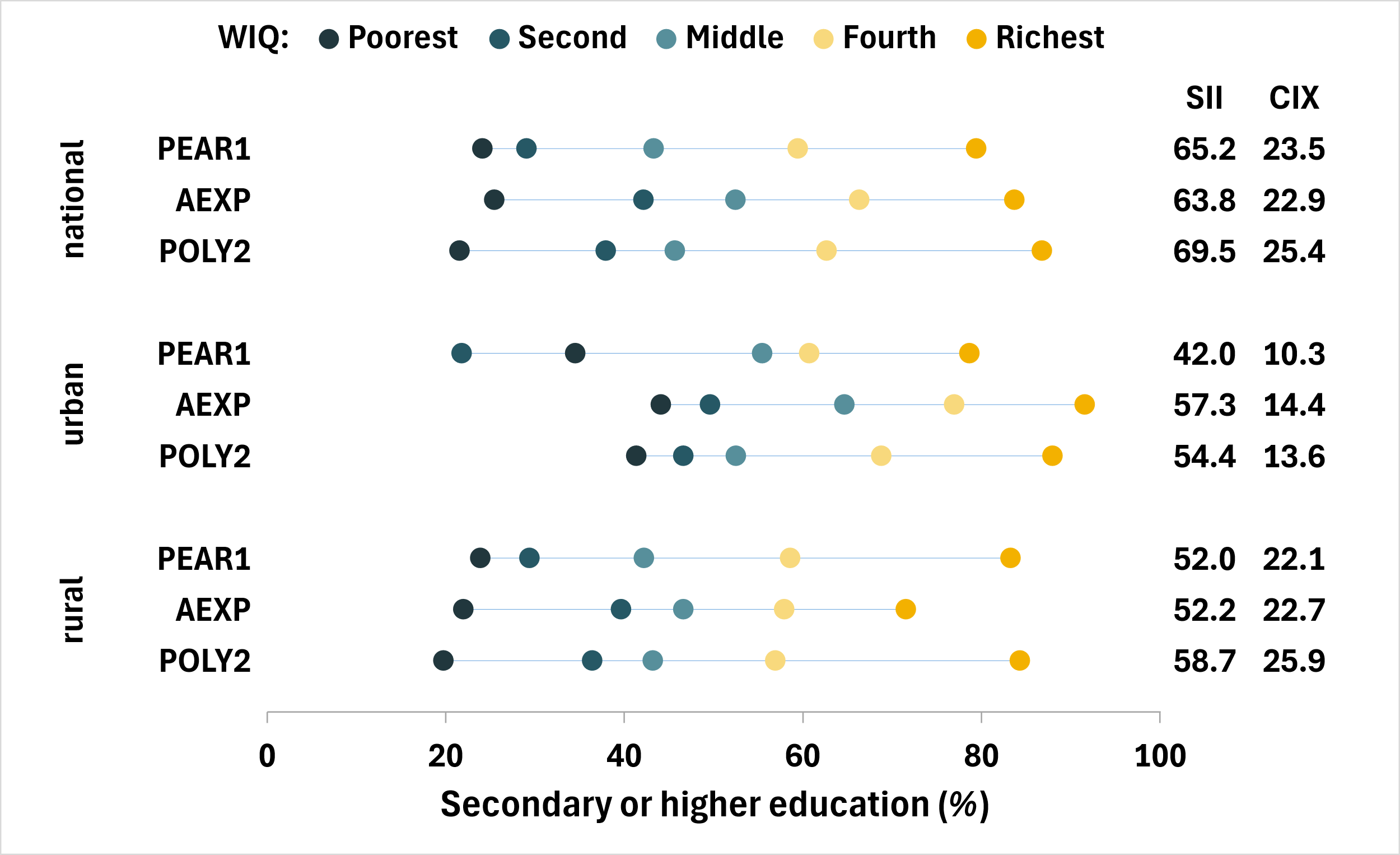

Figure SM21. Proportion of households with at least one member with secondary or higher education by wealth quintile, index, and geographic area – Togo

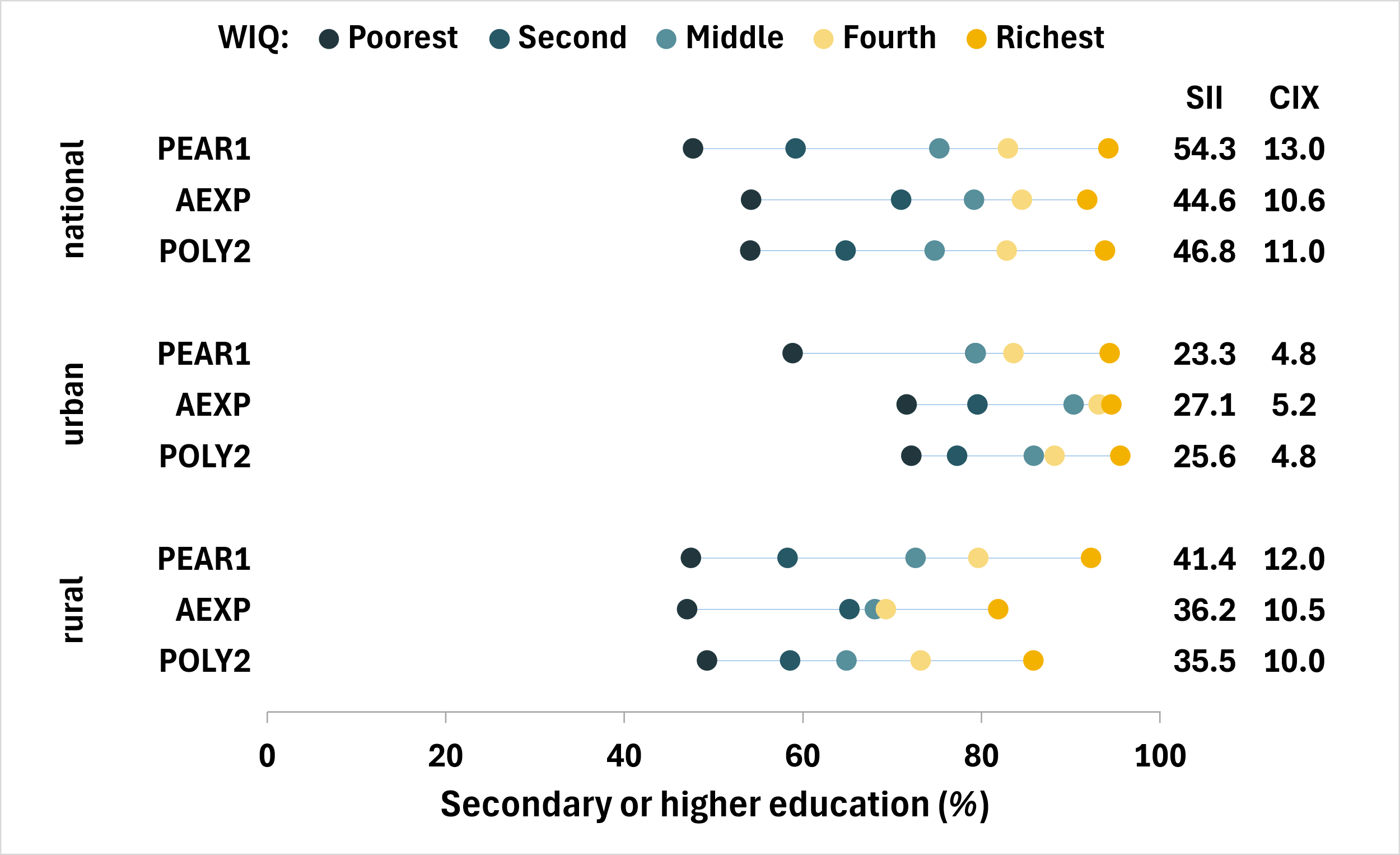

Figure SM22. Proportion of households with at least one member with secondary or higher education by wealth quintile, index, and geographic area – Uganda

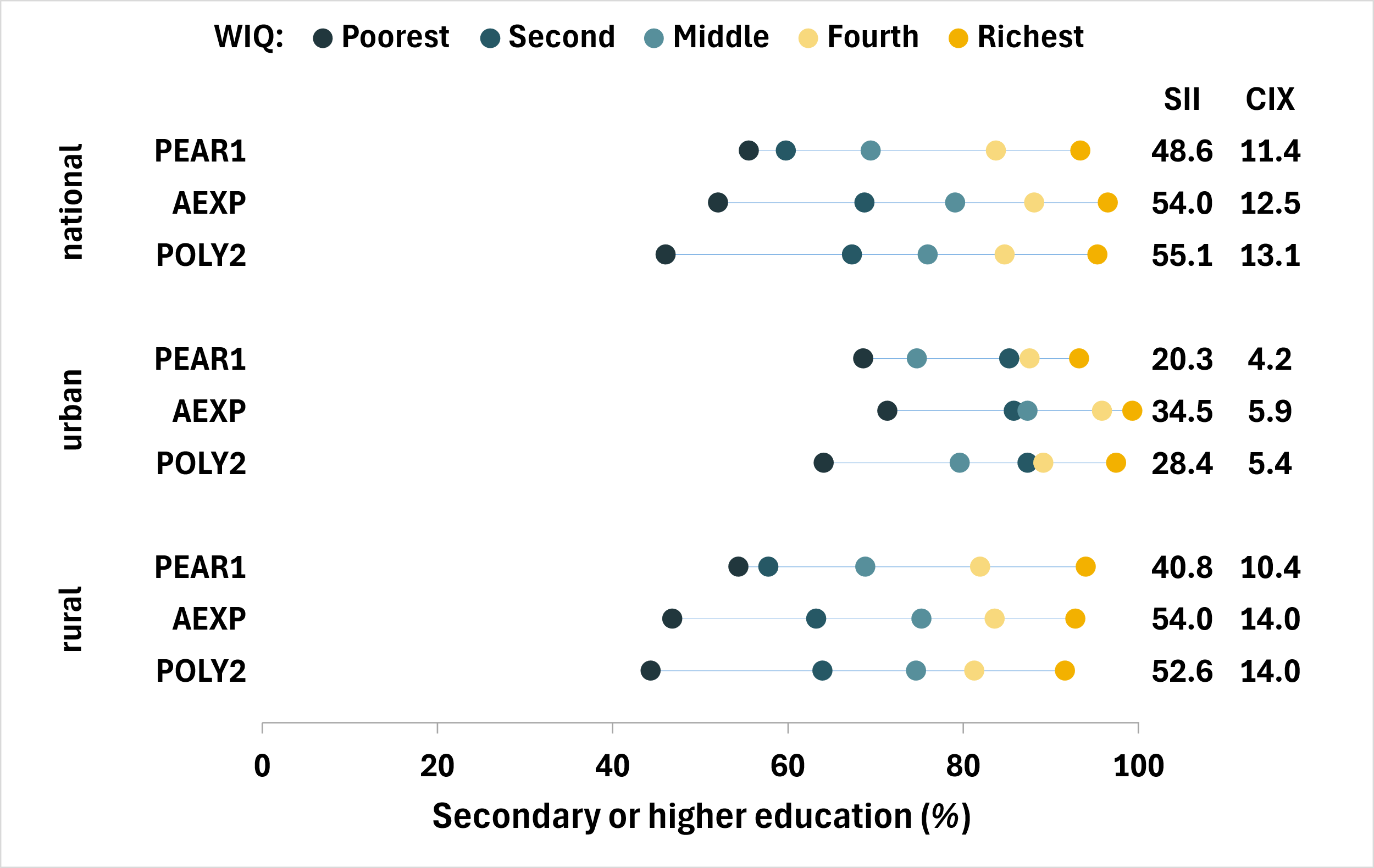

Table SM25. Table showing national summary measures of absolute (SII) and relative (CIX) inequality by index and by country

|  |  |  |  |  |  | **Slope Index of Inequality (SII)** | | |  | **Concentration Index (CIX)** | | |
| --- | --- | --- | --- | --- | --- | --- | --- | --- | --- | --- | --- | --- |
| **ISO** | **Country** | **Survey** | **Source** | **level** |  | **PEAR1** | **AEXP** | **POLY2** |  | **PEAR1** | **AEXP** | **POLY2** |
| BEN | Benin | 2021-22 | LSMS | national |  | 59.3 | 45.9 | 43.7 |  | 17.4 | 13.2 | 12.4 |
| BFA | Burkina Faso | 2021-22 | LSMS | national |  | 56.3 | 49.6 | 49.5 |  | 18.1 | 15.3 | 15.6 |
| CIV | Côte d'Ivoire | 2021-22 | LSMS | national |  | 65.8 | 52.5 | 45.6 |  | 18.9 | 14.5 | 12.5 |
| ETH | Ethiopia | 2021-22 | LSMS | national |  | 70.2 | 36.4 | 71.9 |  | 33.4 | 15.7 | 33.5 |
| GNB | Guinea-Bissau | 2021-22 | LSMS | national |  | 78.8 | 68.6 | 58.7 |  | 39.3 | 33.2 | 27.5 |
| MWI | Malawi | 2019-20 | LSMS | national |  | 59.9 | 58.1 | 50.1 |  | 36.6 | 35.3 | 29.7 |
| MLI | Mali | 2021-22 | LSMS | national |  | 76.7 | 64.6 | 66.8 |  | 46.7 | 38.7 | 40.0 |
| NER | Niger | 2021-22 | LSMS | national |  | 60.0 | 55.5 | 53.2 |  | 27.1 | 24.7 | 22.7 |
| SEN | Senegal | 2021-22 | LSMS | national |  | 48.4 | 45.9 | 47.1 |  | 12.4 | 11.2 | 11.5 |
| TZA | Tanzania | 2021-22 | LSMS | national |  | 65.2 | 63.8 | 69.5 |  | 23.5 | 22.9 | 25.4 |
| TGO | Togo | 2021-22 | LSMS | national |  | 54.3 | 44.6 | 46.8 |  | 13.0 | 10.6 | 11.0 |
| UGA | Uganda | 2019-20 | LSMS | national |  | 48.6 | 54.0 | 55.1 |  | 11.4 | 12.5 | 13.1 |

Table SM26. Table showing urban summary measures of absolute (SII) and relative (CIX) inequality by index and by country

|  |  |  |  |  |  | **Slope Index of Inequality (SII)** | | |  | | **Concentration Index (CIX)** | | |
| --- | --- | --- | --- | --- | --- | --- | --- | --- | --- | --- | --- | --- | --- |
| **ISO** | **Country** | **Survey** | **Source** | **level** |  | **PEAR1** | **AEXP** | **POLY2** | |  | **PEAR1** | **AEXP** | **POLY2** |
| BEN | Benin | 2021-22 | LSMS | urban |  | 52.3 | 44.1 | 43.3 | |  | 13.0 | 10.7 | 10.4 |
| BFA | Burkina Faso | 2021-22 | LSMS | urban |  | 41.8 | 48.2 | 41.8 | |  | 10.4 | 10.5 | 9.7 |
| CIV | Côte d'Ivoire | 2021-22 | LSMS | urban |  | 42.9 | 41.5 | 36.2 | |  | 9.8 | 9.0 | 7.9 |
| ETH | Ethiopia | 2021-22 | LSMS | urban |  | 56.6 | 44.5 | 56.4 | |  | 13.9 | 11.1 | 14.3 |
| GNB | Guinea-Bissau | 2021-22 | LSMS | urban |  | 57.6 | 58.6 | 57.5 | |  | 17.2 | 17.3 | 16.6 |
| MWI | Malawi | 2019-20 | LSMS | urban |  | 60.1 | 62.0 | 60.8 | |  | 20.2 | 20.0 | 18.6 |
| MLI | Mali | 2021-22 | LSMS | urban |  | 70.9 | 64.3 | 71.3 | |  | 22.0 | 19.7 | 22.2 |
| NER | Niger | 2021-22 | LSMS | urban |  | - | 49.3 | 50.6 | |  | - | 12.1 | 11.9 |
| SEN | Senegal | 2021-22 | LSMS | urban |  | 27.9 | 38.9 | 41.4 | |  | 6.5 | 8.1 | 8.6 |
| TZA | Tanzania | 2021-22 | LSMS | urban |  | 42.0 | 57.3 | 54.4 | |  | 10.3 | 14.4 | 13.6 |
| TGO | Togo | 2021-22 | LSMS | urban |  | 23.3 | 27.1 | 25.6 | |  | 4.8 | 5.2 | 4.8 |
| UGA | Uganda | 2019-20 | LSMS | urban |  | 20.3 | 34.5 | 28.4 | |  | 4.2 | 5.9 | 5.4 |

Table SM27. Table showing rural summary measures of absolute (SII) and relative (CIX) inequality by index and by country

|  |  |  |  |  |  | **Slope Index of Inequality (SII)** | | |  | | **Concentration Index (CIX)** | | |
| --- | --- | --- | --- | --- | --- | --- | --- | --- | --- | --- | --- | --- | --- |
| **ISO** | **Country** | **Survey** | **Source** | **level** |  | **PEAR1** | **AEXP** | **POLY2** | |  | **PEAR1** | **AEXP** | **POLY2** |
| BEN | Benin | 2021-22 | LSMS | rural |  | 47.1 | 38.4 | 31.3 | |  | 16.5 | 13.2 | 10.8 |
| BFA | Burkina Faso | 2021-22 | LSMS | rural |  | 31.1 | 31.4 | 33.0 | |  | 11.9 | 11.8 | 12.3 |
| CIV | Côte d'Ivoire | 2021-22 | LSMS | rural |  | 42.3 | 29.6 | 30.9 | |  | 16.5 | 11.5 | 11.7 |
| ETH | Ethiopia | 2021-22 | LSMS | rural |  | 44.6 | 14.9 | 49.1 | |  | 27.6 | 8.7 | 29.6 |
| GNB | Guinea-Bissau | 2021-22 | LSMS | rural |  | 23.4 | 26.6 | 12.9 | |  | 25.9 | 27.4 | 13.4 |
| MWI | Malawi | 2019-20 | LSMS | rural |  | 41.9 | 41.6 | 38.5 | |  | 31.2 | 31.0 | 28.1 |
| MLI | Mali | 2021-22 | LSMS | rural |  | 42.6 | 30.9 | 32.2 | |  | 40.1 | 30.5 | 31.6 |
| NER | Niger | 2021-22 | LSMS | rural |  | 34.3 | 32.4 | 29.8 | |  | 19.1 | 17.9 | 15.1 |
| SEN | Senegal | 2021-22 | LSMS | rural |  | 36.6 | 31.9 | 35.6 | |  | 11.9 | 9.5 | 10.6 |
| TZA | Tanzania | 2021-22 | LSMS | rural |  | 52.0 | 52.2 | 58.7 | |  | 22.1 | 22.7 | 25.9 |
| TGO | Togo | 2021-22 | LSMS | rural |  | 41.4 | 36.2 | 35.5 | |  | 12.0 | 10.5 | 10.0 |
| UGA | Uganda | 2019-20 | LSMS | rural |  | 40.8 | 54.0 | 52.6 | |  | 10.4 | 14.0 | 14.0 |

1.	Filmer D, Pritchett LH. Estimating wealth effects without expenditure data-or tears: An application to educational enrollments in states of India. *Demography*. 2001;38(1):115.
